# Sickness presenteeism due to respiratory infection in the English workforce: prevalence estimates and demographic factors from the Winter COVID-19 Infection Study (WCIS)

**DOI:** 10.64898/2026.02.13.26346245

**Authors:** Mark Gideon Burdon, Sam Denson, Maria Tang, Jonathon Mellor, Thomas Ward

## Abstract

**Background:** Working while sick (presenteeism) with an infectious disease contributes to the spread of infections and is detrimental to productivity. Respiratory illnesses are a common cause of sickness in the working population and understanding the prevalence of presenteeism linked to respiratory illness is therefore important.

**Methods:** Winter Covid Infection Study (WCIS) panel members in work aged 18-64 were surveyed in February - March 2024 and asked about presenteeism in the previous 28 days. Multilevel regression and poststratification was used to estimate the prevalence and length of presenteeism and its effect on productivity in the English workforce, as approximated using the WCIS survey sample calibrated to census proportions. Differences by demographic groups and work sector were also analysed.

**Results:** Around one in six working adults in England worked while sick with a respiratory infection during the study period, and one in ten attended a non-home workplace. Overall, around one day per adult was spent working while sick with a respiratory infection, approximately half of which was non-home working. Respondents felt they were able to work at around three-quarters of their usual capacity while sick. Presenteeism was more common among respondents who were younger, White, worked in a hybrid pattern, lived in larger households, had Long COVID-19, or worked in teaching and education.

**Conclusion:** Working while sick with a respiratory infection is relatively common, including among those who primarily work away from the home.

**Key messages:** Around one in six working-age adults in employment worked while sick with a respiratory infection during the study period (Feb-Mar 2024). - The likelihood of working while sick with a respiratory infection varied by demographic group and work sector. - On average, survey respondents said they could work at around three quarters their normal effectiveness while sick with a respiratory infection.

## Introduction

Presenteeism - presenting for work despite being sick or otherwise in poor health - is approximately as common in the working population as sickness absence^1^. It has long been recognised as detrimental to economic productivity^2^ and increases the spread of infectious disease^3^. Presenteeism is therefore of interest to policymakers both for economic and public health reasons. Previous research has estimated prevalence of presenteeism in various occupational settings and countries, estimated risk factors contributing to higher presenteeism rates in some groups and used qualitative methods to explore reasons why people worked while sick^4,5^. A higher risk of presenteeism has previously been found to be associated with work-related factors such as long working hours^1^, personal characteristics such as disability or sex^6^, and the broader economic context^7^.

This study uses responses from the Winter COVID-19 Infection Study (WCIS)^8,9^ run by the UK Health Security Agency (UKHSA) and Office for National Statistics (ONS) to estimate the prevalence of presenteeism while suffering a respiratory infection in February and March of 2024 among working adults aged 18-64 in England. Group-level estimates are also made for a range of demographic groups and work sectors. These estimates are used to explore the factors associated with presenteeism, and produce estimates of the potential total productivity loss in each sector of the economy due to working while sick with a respiratory infection. Although many studies have explored risk factors for presenteeism, presenteeism prevalence estimates for a whole population using a representative sample are scarce (e.g.^10^ and^11^; for literature reviews, see^4^ and^5^). This study therefore provides valuable insight into the potential economic and health implications of presenteeism driven by seasonal respiratory infections.

## Methods

### Data and variables

Between November 2023 and March 2024, the WCIS produced fortnightly estimates of the community prevalence of COVID-19. The panel consisted of previous participants in the COVID-19 Infection Survey, which launched in England in April 2020 and was based on a random sample of households^12^. Between the 12th of February 2024 and the 7th March 2024, respondents were asked questions about respiratory infections and work. 24,394 WCIS respondents in work aged 18-64 in England reported the number of days in the last 28, if any, they had worked while sick with a respiratory infection (presenteeism). For those who reported having had a respiratory infection, they were also asked how many days they worked in the previous 28; how many days they worked from home while sick; and how able they were to complete work tasks while sick on a 0-10 scale.

Because of differential non-response to the invitation to participate in WCIS among different demographics, the sample is not representative of the English working population. Poststratification was used to make the survey sample match census population proportions for age group, sex, ethnicity and work sector; details on how WCIS variables were processed to align with census categories and allow poststratification are given in the Supplementary Materials. Bias in dimensions other than those four variables remains possible.

Respondents who did not report any sickness due to respiratory infection were not asked about their working patterns, and therefore estimating the proportion of workdays affected by presenteeism required addressing this omission. A range of likely values for the number of days worked for each demographic combination were generated by multiple imputation using Random Forests via the *mice* R package^13^. The full range of uncertainty from the imputation may not be propagated into the results, and so the resulting credible intervals should be treated with caution. The model was then estimated across all fifteen imputed datasets and the results pooled.

## Models

### Models 1-2: What proportion of working adults worked while sick, in total and outside their home?

To estimate the proportion of working adults who worked while sick with a respiratory infection (presenteeism) in total and outside their home, we estimated Bayesian multi-level binomial regression models where the response variable was a binary indicator representing whether the respondent reported working while sick at any point in the last 28 days, either in total (in Model 1, M1) or outside the home (in Model 2, M2). Because choices around presenteeism are likely to be correlated with respondent demographics, predictors for variables for age group, sex, ethnicity (collapsed), IMD decile, number of days of sickness absence, working location, SARS-CoV-2 LFT positivity, self-reported long COVID, household size, and region were added. The model also includes random intercepts by work sector and random slopes for sex within work sector. These random slopes allow the relationship between sex and presenteeism to vary in different parts of the economy, as sex differences in potentially relevant unmeasured dimensions such as contractual status, wellbeing and pay may potentially differ by work sector^14,15^.

### Models 3-5: How many days on average did working adults work while sick with a respiratory infection, in total and outside their home? How does this compare to sickness absence?

Model 3 (M3) was used to estimate the number of days of presenteeism while sick with a respiratory infection. Because of the additional risk of onward transmission from working outside the home the number of days reported working while sick with a respiratory infection outside the home was also estimated (in Model 4, M4). Because in each case the response variable is an overdispersed count with many zeros, two-part (hurdle negative binomial) models were used. The same set of controls were used as in M1 and M2, for both the hurdle and conditional components of M3 and M4. This was based on the expectation that the same factors that affect whether any presenteeism occurs may also affect the length of presenteeism.

The total number of days worked outside the home while sick, as estimated by M4, is important because of the potential for onward transmission of respiratory infections. However, for comparison between different groups where average working patterns vary, estimating presenteeism outside the home as a proportion of days worked is preferable. In Model 5 (M5), the proportion of workdays that were worked outside the home while sick is directly estimated using a Bayesian ordered Beta regression model, based on partly imputed data for the number of days worked and using the same set of control variables.

### Model 6: What was the self-reported impact of presenteeism on productivity?

Model 6 (M6) estimates respondents’ self-reported ability to work while sick with a respiratory infection. WCIS respondents who reported working while sick with a respiratory infection were asked to rate how able they were to work while sick on a Likert scale of zero to ten, where ten is their normal ability when healthy. This variable was transformed to a ratio and modelled using ordered Beta regression, with the same set of predictors as the other models. Only respondents who reported presenteeism with a respiratory infection in the 28-day study period were included. The model allows comparison of self-reported ability to work across demographic groups.

The ordered Beta regression model for M6 takes the same structure as M5, but in this case models self- reported ability to work as a proportion between 0 and 1.

### Implementation and Priors

Binomial and negative-binomial models were estimated in the *brms* package^16^ and ordered Beta regression models in the *ordbetareg* package^17^, sampled with four chains and 4000 iterations per chain, except for M5 which had 6000 iterations per chain to increase the effective sample size. Across all models, weakly informative priors were used to help convergence and weakly regularize effect sizes. Further model specification details, and visualisations of the priors, are presented in the Supplementary Materials. To mitigate differences between the WCIS survey sample and known proportions, all estimates are poststratified by age group, sex, ethnicity and work sector.

## Results

### What proportion of working adults worked while sick with a respiratory infection, in total and outside the home (M1 and M2)?

As Figure 1 shows, respondents were more likely to work while sick if they also took time off work for sickness, had taken a positive LFT for SARS-CoV-2, had Long COVID-19, were of White ethnicity, or worked in a hybrid arrangement. Larger household sizes also increased the probability of working while sick. Respondents were less likely to work while sick if they were aged between 55 and 64. Poststratified average marginal effects that isolate the estimated effect of each variable are shown in the Supplementary Materials.

**Figure 1.**
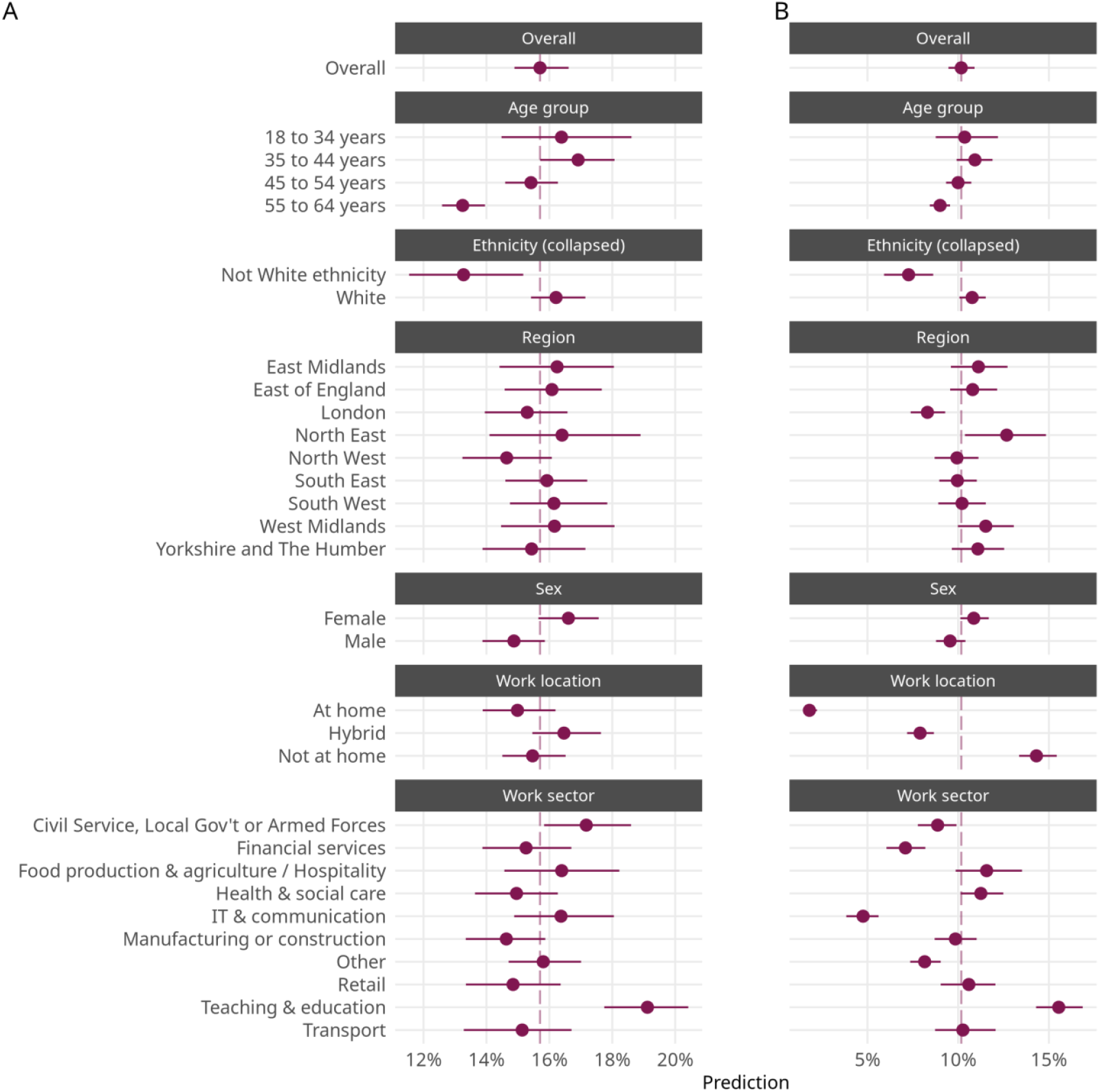
Average estimated proportions and 95% credible intervals of the working population who reported working while sick with a respiratory infection (in total or outside the home) during the study period. Estimates are posterior predictions from the Bayesian logistic regression models M1 and M2, after poststratification, and are averaged within demographic groups. A: Results from M1 (total presenteeism). B: Results from M2 (presenteeism outside the home).

Factors affecting the probability of working while sick outside the home were similar and in the same direction, but also reflected differences in usual working locations. As expected, individuals who normally work outside the home or on a hybrid basis were more likely to work while sick outside the home.

Overall, 15.70% (95% CrI: 14.89%, 16.61%) of the working population were estimated to have worked while sick with a respiratory infection during the study period in total, after poststratification. 10.17% (95% CrI: 9.47%, 10.90%) were estimated to have worked outside the home while sick with a respiratory infection.

### How often did working adults work while sick with a respiratory infection, in total and outside their home (M3, M4 and M5)?

As demonstrated by Figure 2, presenteeism days were positively associated with length of sickness absence, having Long COVID-19 and having a positive SARS-CoV-2 LFT. Larger household sizes, being white, the 45-54 age group and being male had weaker positive associations with presenteeism days. Note that this model does not take into account how many days were worked during the study period. For comparison, M3b in the Supplementary Materials estimates the number of days of sickness absence.

**Figure 2.**
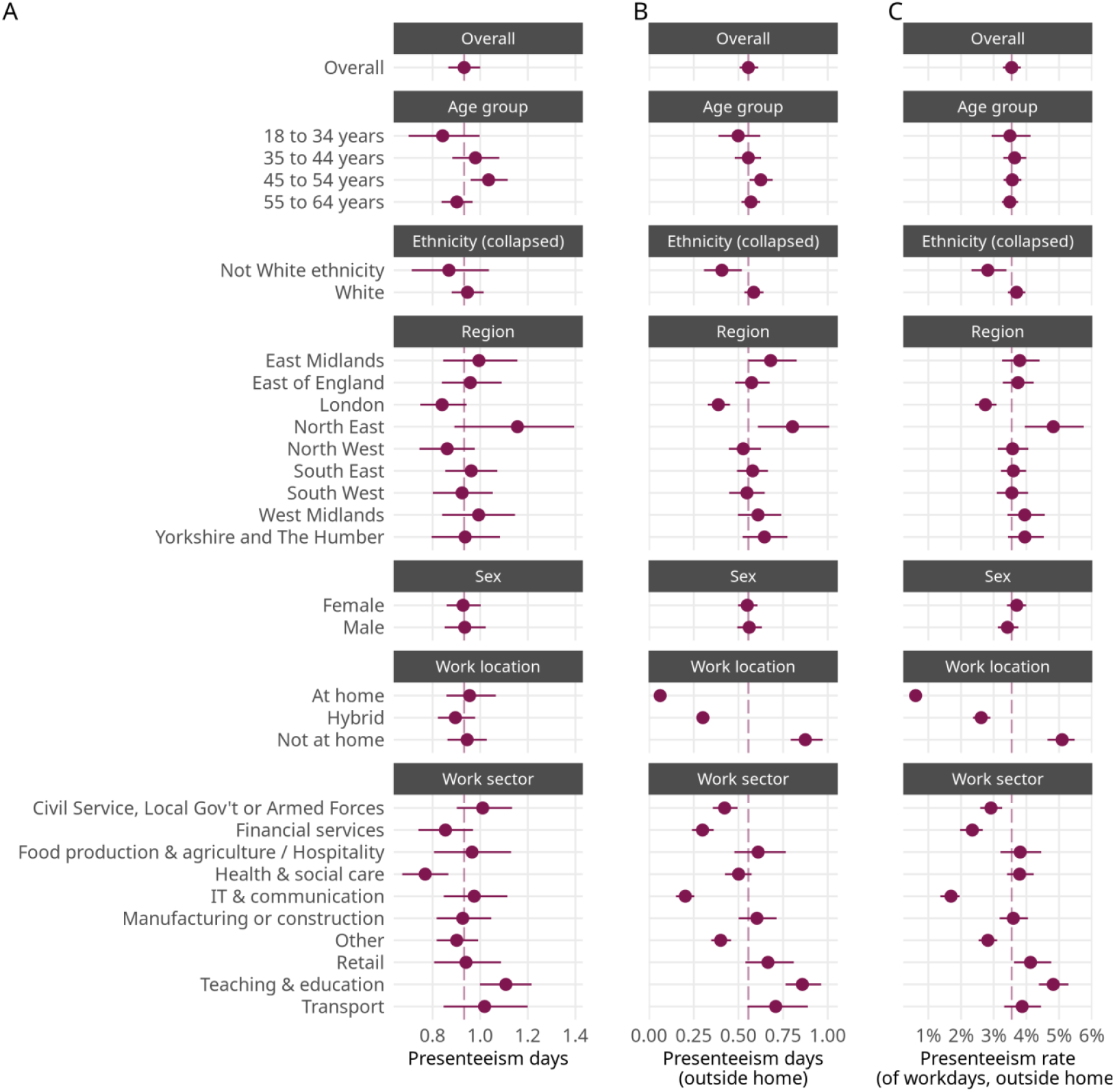
Average estimated number or rate of days worked while sick with a respiratory infection during the study period (A: presenteeism days, B: presenteeism days outside the home, C: rate of presenteeism days outside the home), with 95% credible intervals. Predictions are derived from the Bayesian hurdle negative binomial regression models 3-5 after poststratification, and are averaged within demographic groups.

Presenteeism outside the home had the same main risk factors, but was also positively associated with the 45-54 age group, and with hybrid or non-home working, as expected.

On average, working-age adults in work were estimated to have worked while sick with a respiratory infection in any location for 0.93 days in the 28-day study period, after poststratification (95% CrI: 0.87, 1.00 days). They were estimated to have worked while sick with a respiratory infection outside the home for 0.56 days (95% CrI: 0.51, 0.61 days).

M5 estimated that people worked while sick with a respiratory infection in 3.5% of days worked in the 28-day study period, after poststratification (95% CrI: 3.3%, 3.8%).

### What was the self-reported impact of presenteeism on productivity (M6)?

On average, people who reported being sick with a respiratory infection said that their ability to work while sick with a respiratory infection was 76.8% of their usual ability (95% CrI: 75.8%, 77.8%). On average, respondents in age groups over 45 rated their ability to work while sick higher than the youngest category. Respondents who were male, White or worked outside the home also rated their ability to work while sick higher than others. Lower average ability to work while sick was reported by respondents when they reported Long COVID-19, took longer sickness absence, who worked in teaching and education. However, these average differences were relatively minor: the highest positive effect size estimate was approximately 4% (for the age group 55-64) and the lowest negative effect size estimate was approximately minus 6% (for Long COVID-19). These estimates are shown in Figure 3.

**Figure 3.**
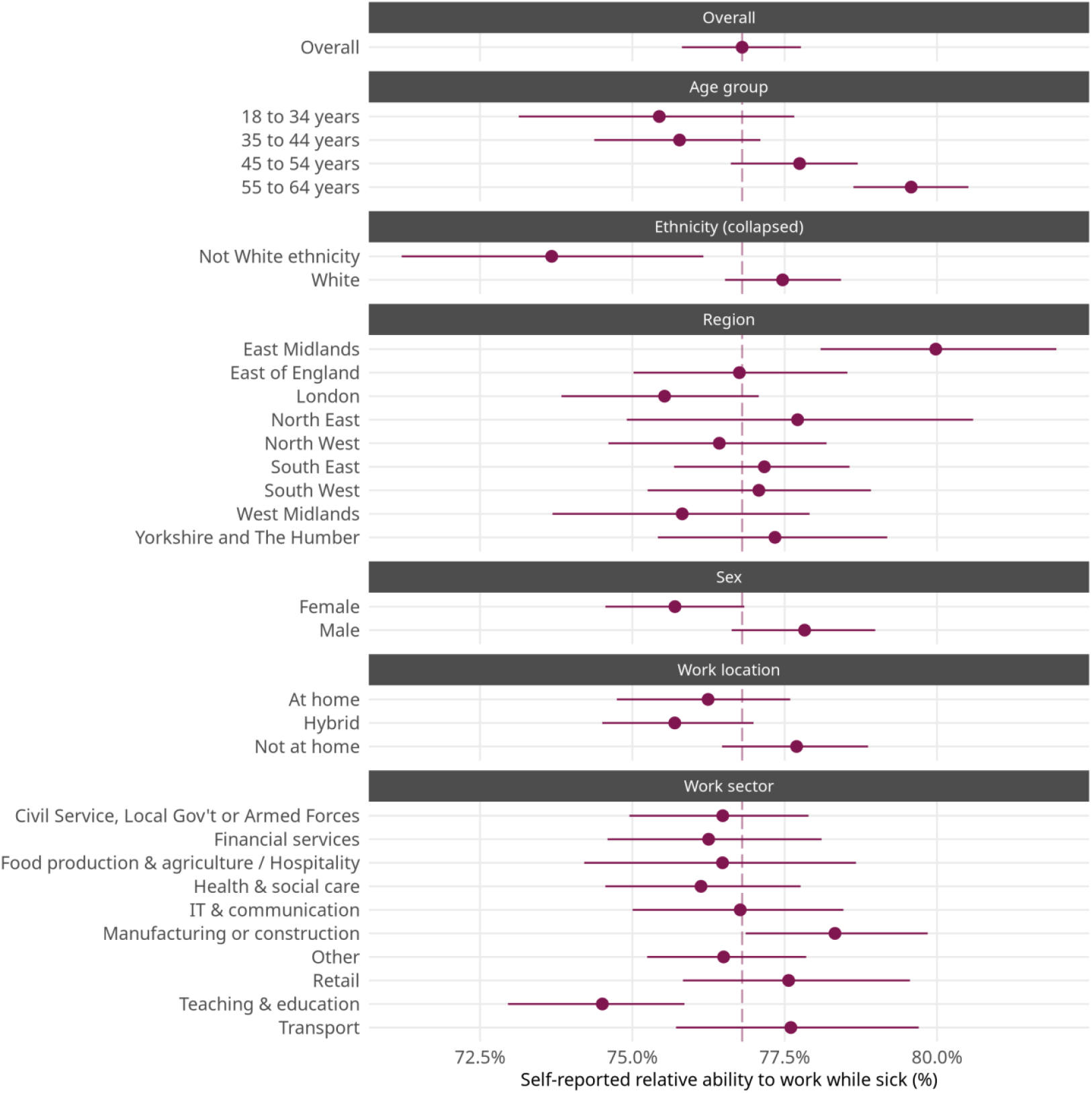
Average estimated self-rated relative ability to work while sick with a respiratory infection, with 95% credible intervals. Predictions were derived from the Bayesian ordered beta regression model M6, after poststratification, and are averaged within demographic groups. 100% represents the respondent’s usual productivity, and only respondents who reported being sick with a respiratory infection were included

Figure 4 shows the average number of productivity days lost per person in each work sector during the 28-day study period. This was calculated by multiplying poststratified estimated number of days worked while sick with a respiratory infection from M3 with the poststratified estimated self-reported ability to work while sick with a respiratory infection from M6; for example, two days of working while sick at 50% effectiveness would equal a loss of one productivity day. These estimates are then multiplied by the number of people working in each work sector from the 2021 Census and divided by 365.25 to create a “years of productivity lost” metric, which is also shown in Figure 4 and gives an idea of the total scale of respiratory illness’ impact on in-work productivity in different work sectors during the 28-day study period. Note that this assumes the counterfactual is that the individuals would be working while fully well, and so this model is looking at the effect of the illness; absenteeism would result in 0% effectiveness. Estimating workplace transmission or modelling the decision to work or not is beyond the scope of this paper and the suitability of the data.

**Figure 4.**
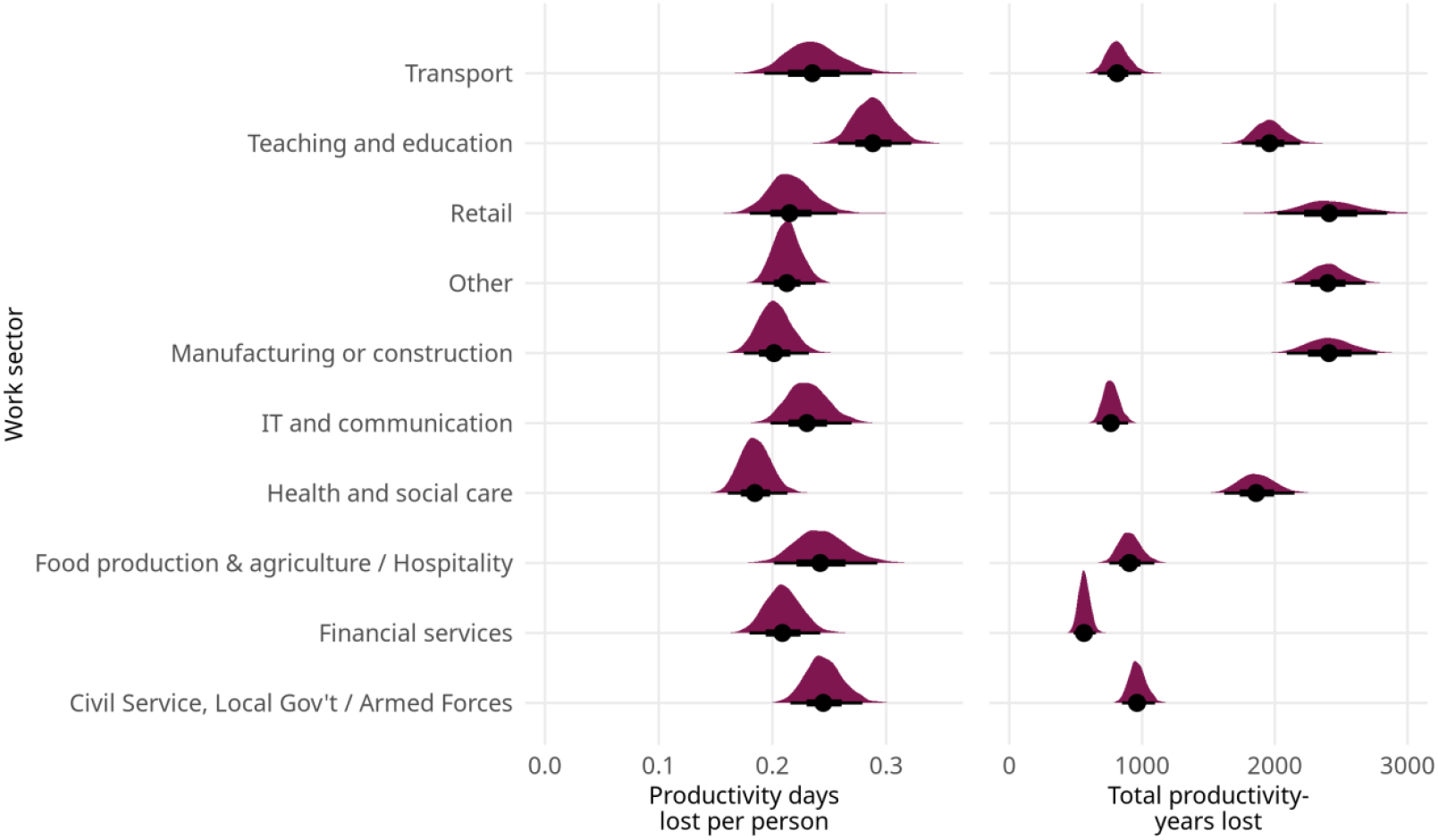
Probability density plots showing the estimated number of productivity days lost per person and total productivity-years lost by work sector. These estimates were produced by combining survey respondents’ self-reported effectiveness ratings from M6 with the estimated numbers of days of presenteeism from M3, and in the case of total productivity-years, by multiplying this by the number of people working in that work sector.

## Discussion

Overall, working while sick with a respiratory infection was relatively common in this survey study, with approximately one in six working adults in England working while sick with a respiratory infection at some point during a 28-day window, and one-in-ten working while sick outside the home. Around one day per month per working adult was spent working while sick, approximately half of which was outside the home. Since survey respondents rated their ability to work while sick as being around three-quarters of their usual capability, this represents a significant economic impact as well as potentially a meaningful contributor to the spread of respiratory infections in the population. However, we would expect several winter viruses to be prevalent in February and March^18^, so this cannot be simply extrapolated to the rest of the year.

After adjusting for other factors, people of White ethnicity were estimated to be more likely to work while sick, but gave higher self-evaluations of their ability to work while sick. However, ethnicity had no clear effect on the number of days of presenteeism, suggesting that White respondents may have been more likely to engage in presenteeism for short durations. This could also explain this group’s higher self-reported impact of presenteeism on productivity for White individuals, if shorter periods of presenteeism are perceived to have less of an impact. Related research shows some evidence of ethnic minority populations in the UK to be of higher risk of severe outcomes from influenza^19,20^, have lower access to care^21^, and lower job control within the same work sector (influence over what work you do and how you do it)^22^, which could affect presenteeism.

There was also strong evidence that age was associated with presenteeism after controlling for covariates, with the oldest age group (55-64) less likely to work while sick and reporting a higher negative association between presenteeism and productivity, compared to the youngest age group (18-34). This could result from differences in health vulnerability or societal and workplace expectations. The 45-54 age group also reported lower productivity compared to the youngest age group, but spent more days working while sick outside the home. These patterns may reflect age-related differences in perceived pressure to work while sick, influenced by factors such as seniority, workplace culture, or financial responsibilities. Financial responsibilities may similarly play a role in higher presenteeism levels for larger households, although larger households also have a higher infection risk, as having more household members increases the likelihood of infection being brought into the house and of its transmission to other household members. Existing findings on age and general presenteeism are mixed^4,5^.

Across work location types, hybrid workers were slightly more likely to work while sick, but no meaningful difference was found in their total days of presenteeism after adjusting for other factors. This runs contrary to expectations from the literature, where home workers are expected to work while sick more often because of the lack of commute and risk of onward infection^23^. Hybrid workers lay about halfway between remote and on-site workers in terms of likelihood of occurrence and proportion of days of presenteeism outside the home, but closer to remote workers in terms of number of days. Several factors could contribute to this, including the ratio of remote to on-site days for hybrid workers, whether they have the flexibility to choose their work location on a given day when sick, and the work culture across different work locations. The self-reported relative ability to work while sick was slightly worse for on-site workers, perhaps due to the nature of on-site job or the added strain of a commute and in-person activities.

Overall rates of presenteeism and total presenteeism days were similar across work sectors, but greater differences emerged when considering only presenteeism outside the home, likely due to sector-wide differences in work location. Workers in teaching and education are more likely to work while sick than other workers, both in general and outside the home. Further, self-reported relative ability to work while sick is lowest in teaching and education. European studies also found that education staff have higher presenteeism^24^, as well as those in the agriculture and health sectors^25^. These differences may be influenced by work-related factors that correlate with sector, but are not measured directly in our survey; for example, stress-related burnout due to excessive job demands can lead to presenteeism^26^.

One limitation of this study is the difficulty in distinguishing between increased risk of illness and increased risk of presenteeism. In an attempt to account for different levels of baseline illness, the models controlled for days of sickness leave during the study period, Long COVID-19 status, and testing positive for SARS-CoV-2. Although for some workers, the decision between working while sick and taking sickness absence will be the result of a decision-making process^27^, there was a positive association between absenteeism and presenteeism, indicating that on the whole one does not displace the other. In addition, the estimates for the rate of presenteeism are dependent on the accuracy of the chosen missing data imputation method. The survey did not capture the number of days worked for respondents without a respiratory infection, and this is therefore MNAR (missing not at random), and we have to rely on the assumption that the number of days worked by those without a respiratory infection can be extrapolated from data on those with one.

Other limitations include that the data entirely derives from a single set of self-reported survey questions, making it vulnerable to social desirability bias and common method bias^28^, and that simplifying assumptions have been made e.g. about the concordance between WCIS and Census work sector classifications.

## Data Availability

Access to protected data is always strictly controlled using legally binding data sharing contracts. UKHSA welcomes data applications from organisations looking to use protected data for public health purposes. To request an application pack or discuss a request for UKHSA data you would like to submit, contact.
A synthetic dataset has been generated and made available that has very similar statistical relationships to those that exist in the real survey data. This allows replication of the analysis without disclosing individual survey responses.

http://github.com/burdonmark/presenteeism

## Funding

This research was not linked to any specific funding. Sam Denson is employed by the Department for Health and Social Care in England. The views expressed are those of the authors and not necessarily those of the Department of Health and Social Care.

## Conflicts of Interest

None declared.

## Data Availability Statement

A synthetic dataset has been generated and made available that has very similar statistical relationships to those that exist in the real survey data. This allows reproduction of the paper’s analysis without disclosing individual survey responses. Code and synthetic data can be found at https://github.com/burdonmark/presenteeism/

UKHSA operates a robust governance process for applying to access protected data that considers: - the benefits and risks of how the data will be used - compliance with policy, regulatory and ethical obligations - data minimisation • how the confidentiality, integrity, and availability will be maintained - retention, archival, and disposal requirements - best practice for protecting data, including the application of ‘privacy by design and by default’, emerging privacy conserving technologies and contractual controls

Access to protected data is always strictly controlled using legally binding data sharing contracts. UKHSA welcomes data applications from organisations looking to use protected data for public health purposes. To request an application pack or discuss a request for UKHSA data you would like to submit, contact.

## Notes

### Competing Interest Statement

The authors have declared no competing interest.

### Clinical Protocols

https://www.ndm.ox.ac.uk/covid-19/covid-19-infection-survey/protocol-and-information-sheets

### Funding Statement

The study did not receive any funding.

### Author Declarations

The study received ethical approval from the South Central Berkshire B Research Ethics Committee (20/SC/0195)

