## Supplementary Materials for "Sickness presenteeism due to respiratory infection in the English workforce: prevalence estimates and demographic factors from the Winter COVID-19 Infection Study (WCIS)"

Presenteeism modelling supplementary materials

### Appendix

#### Data and variables

##### Work sectors

Supplementary Table 1 shows how WCIS work sector categories were mapped to ONS Standard Industrial Classifications (SIC) and which were ultimately combined.

Supplementary Table 1: Relationship between original WCIS categories, ONS Standard Industrial Classification (SIC) and final collapsed work sector categories

| WCIS category | Census SIC | Final category |
| --- | --- | --- |
| None | Does not apply | None |
| Manufacturing or construction | F Construction | Manufacturing or construction |
| Manufacturing or construction | B Mining and quarrying | Manufacturing or construction |
| Manufacturing or construction | C Manufacturing | Manufacturing or construction |
| Manufacturing or construction | D Electricity, gas, air cond supply | Manufacturing or construction |
| Manufacturing or construction | E Water supply, sewerage, waste | Manufacturing or construction |
| Civil Service or Local Government | O Public admin and defence | Civil Service or Local Gov’t & Armed forces |
| Other employment sector | M Prof, scientific, technical activ. | Other employment sector |
| Other employment sector | U Extraterritorial organisations | RSTU Other |
| Other employment sector | T Households as employers | RSTU Other |
| Other employment sector | N Admin and support services | Other employment sector |
| Hospitality - for example hotels or restaurants or cafe | I Accommodation and food services | Hospitality - for example hotels or restaurants or cafe |
| Healthcare | Q Health and social work | Healthcare & social care |
| Financial services. This includes insurance | K Financial and insurance activities | Financial services. This includes insurance |
| Arts or entertainment or recreation | R Arts, entertainment and recreation | RSTU Other |
| Retail sector. This includes wholesale | S Other service activities | RSTU Other |
| Retail sector. This includes wholesale | L Real estate activities | Retail sector. This includes wholesale |
| Retail sector. This includes wholesale | G Wholesale, retail, repair of vehicles | Retail sector. This includes wholesale |
| Information technology and communication | J Information and communication | Information technology and communication |
| Teaching and education | P Education | Teaching and education |
| Food production and agriculture. This includes farming | A Agriculture, forestry and fishing | Food production and agriculture. This includes farming |
| Personal Services - for example hairdressers or tattooists | S Other service activities | RSTU Other |
| Transport. This includes storage and logistics | H Transport and storage | Transport. This includes storage and logistics |
| Social Care | Q Health and social work | Healthcare & social care |
| Armed forces | O Public admin and defence | Civil Service or Local Gov’t & Armed forces |

##### Missing data imputation

Age, sex, work location, region, whether the respondent was white, work sector, household size, IMD decile, number of sickness days and long COVID-19 status were used to impute number of days worked. Fifteen possible values were imputed for each individual using a Random Forest model. Upper and lower limits were imposed to prevent an individual from having more than 28 days of work or sickness absence, or from imputing fewer days of work than the reported number of days worked while sick. The model was then estimated across all fifteen imputed datasets and the results pooled.

As Supplementary Fig. 1 shows, respondents with a respiratory infection differed by up to 5% on some demographic categories - in particular, they were more likely to be female, work in teaching and education, live in larger households, work outside the home and be in the youngest two age categories (under 44 years of age). These differences are likely also correlated with working patterns.

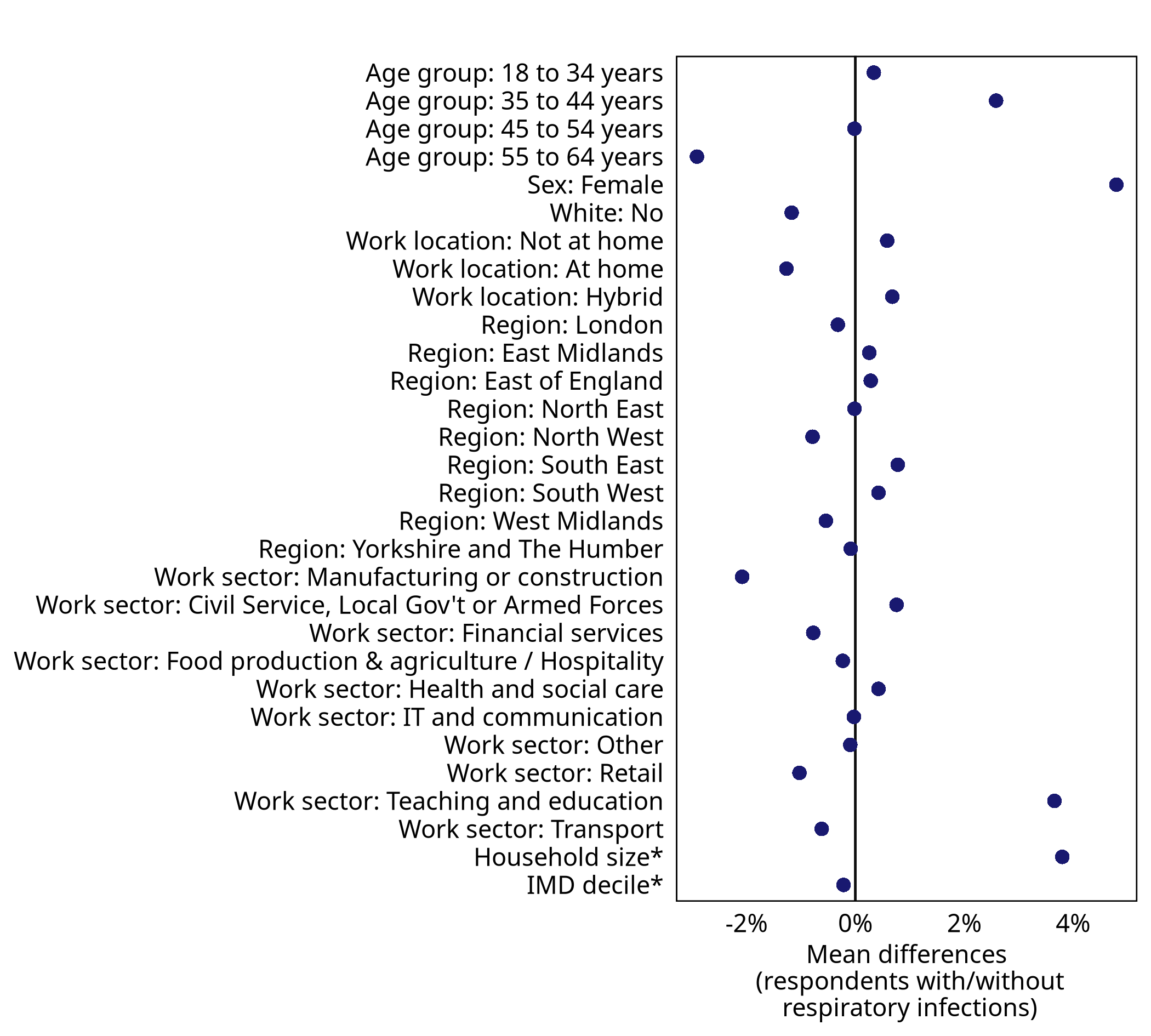

Supplementary Figure 1: Balance plot showing the mean demographic differences between WCIS respondents who reported having a respiratory infection and those who did not. Variables with an asterisk are shown in terms of standardised difference

Summary statistics from imputed data compared with the original data (after excluding missing rows), are shown in Supplementary Table 2.

Supplementary Table 2: Summary statistics showing the difference between number of days worked in the original and imputed data

| Method | Median | Mean | St. Dev |
| --- | --- | --- | --- |
| Imputed via Random Forest | 18.00 | 16.70 | 5.92 |
| Original data | 18.00 | 16.43 | 6.05 |

Supplementary Fig. 2 shows the distribution of number of days worked in the post-imputation data using KDE density plots. As Supplementary Table 2 and Supplementary Fig. 2 show, imputation resulted in very similar distributions of workdays between respondents who reported sickness (original data) and those who did not (imputed data). Non-sick respondents were estimated to be much more likely to work 20 days in the study period than those who were sick.

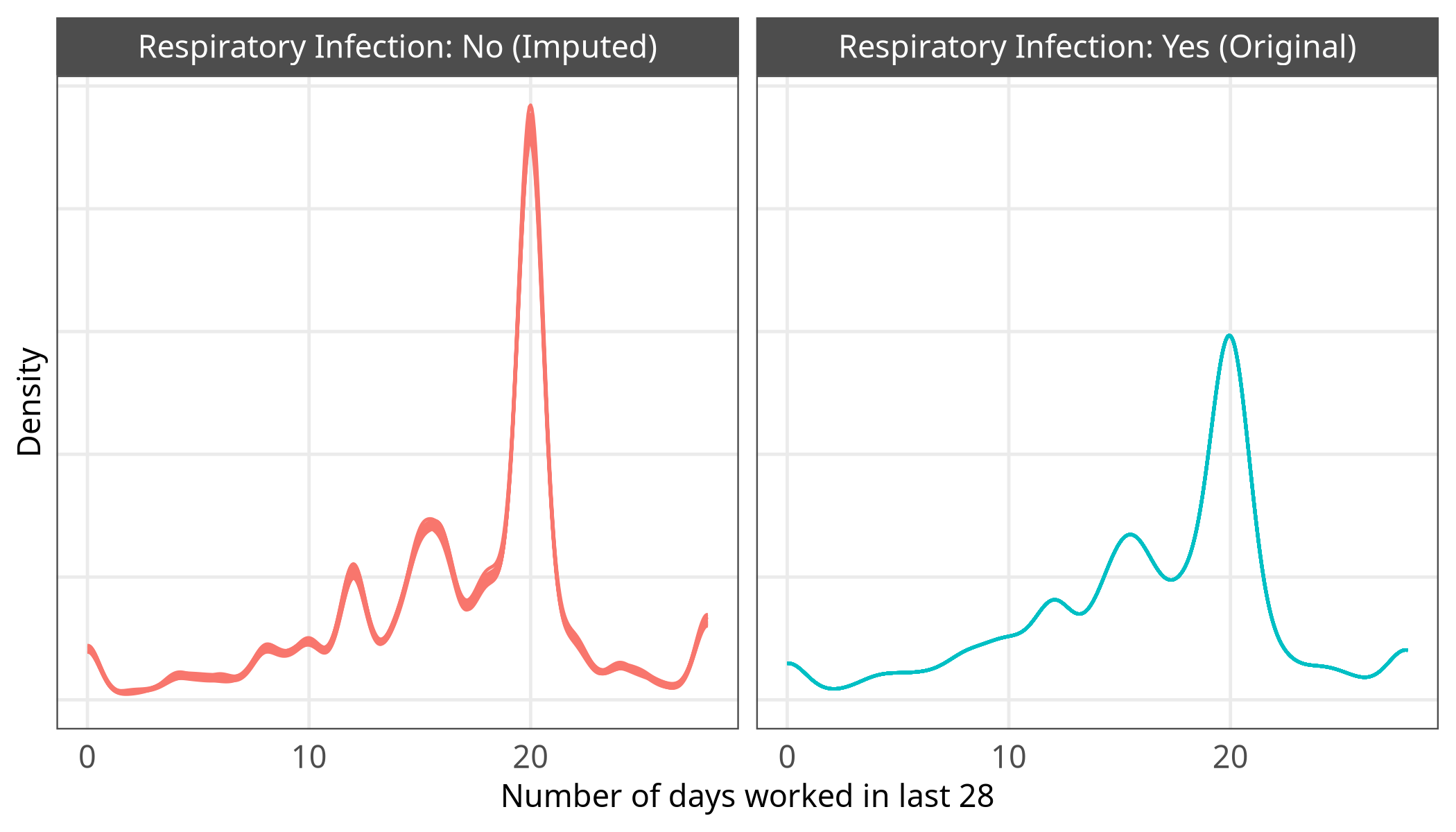

Supplementary Figure 2: Density plot showing the variation in imputed number of workdays among individuals who did not report being sick during the study period, and providing a comparison to real data from individuals who reported a respiratory infection. In the imputed data, each iteration has its own line.

#### Models

##### Variable coding and poststratification

For the poststratification of the estimates by work sector, the categories used in the WCIS survey were aligned with the nearest suitable category in the ONS Census Standard Industrial Classification (SIC). Because of how the SIC categories are constructed, the “Armed forces” and “Civil Service or Local Government” categories were combined into one, “Healthcare” and “Social Care” were combined into “Healthcare & Social care”, and the sectors lettered R S T and U were combined into “Other employment sector”. In addition, to avoid empty cells in the poststratification table, two further categories (“Food production and agriculture. This includes farming” and “Hospitality - for example hotels or restaurants or cafe”) were combined, and ethnicity was collapsed into a White binary variable.

Supplementary Fig. 3 shows that 18-34 year olds and retail or manufacturing workers are the most under-represented by WCIS, while 55-64 year olds and people working in teaching and education are the most over-represented.

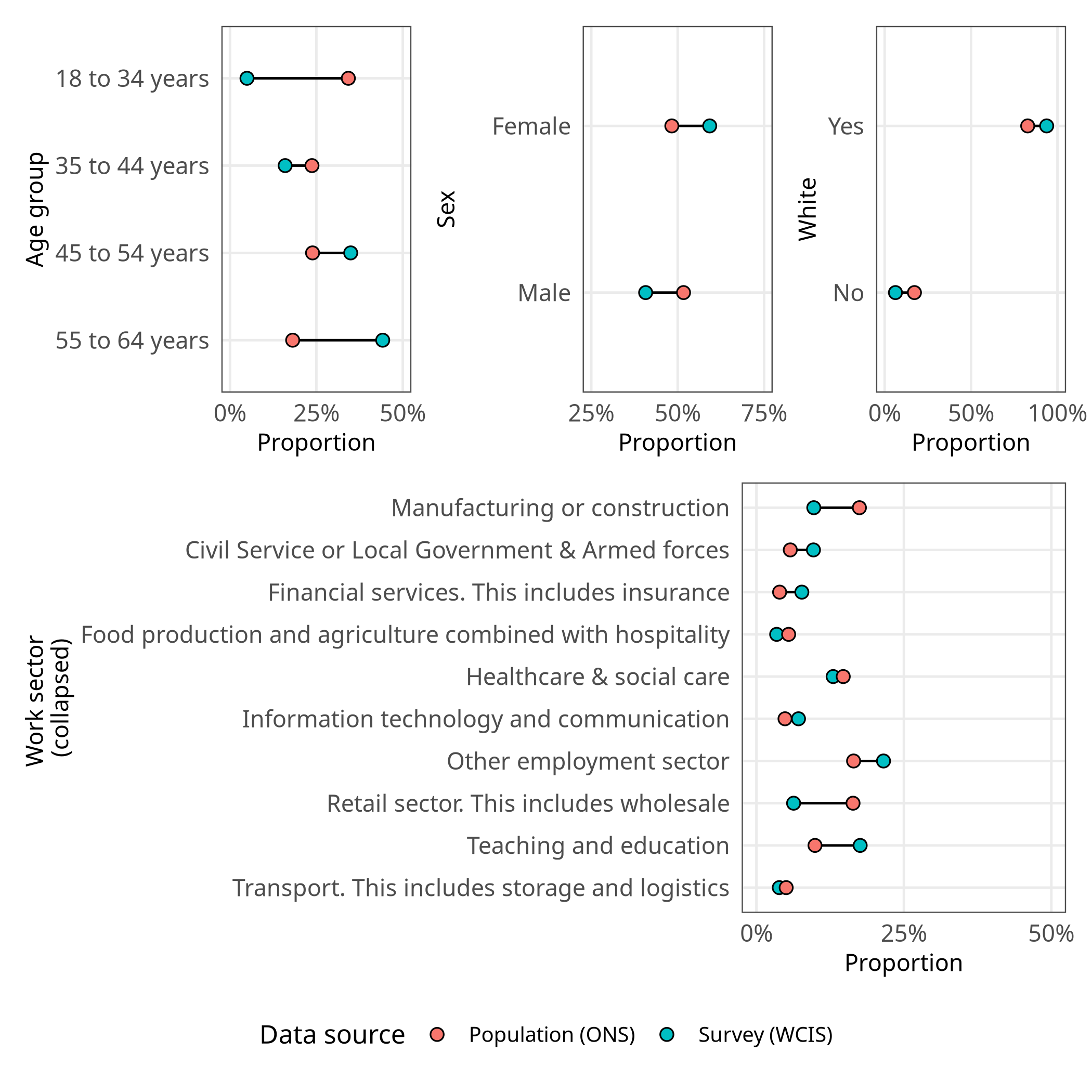

Supplementary Figure 3: Lollipop plots showing the relative proportions of different demographic groups within the WCIS survey respondents, compared to ONS census totals. Compares proporton for age group, sex, ethnicity (collapsed) and work sector (collapsed).

The poststratification frame was constructed using 2021 ONS Census joint distributions of age, sex, White/non-White indicator and collapsed work sector. Because we lack joint distributions for the other variables (e.g. household size), other variables are assumed to follow the same joint distribution in the population as in the survey. The poststratification of each demographic group $i$, where $N$ is the number of individuals and $p$ is the estimated parameter (e.g. proportion of presenteeism) is shown in Equation 1.

$$\frac{\sum_{i} N_{i}p_{i}}{\sum_{i} N_{i}} \left( 1 \right)$$

##### Further model details and formulae

Models M1 and M2 estimate whether the individual $i$ within work sector $j$ worked while sick ($y_{i}$) as arising from a Bernouilli distribution.The probability of presenteeism ($p$) is modelled as the sum of a global intercept $\alpha$, fixed effects for coefficients ($\beta_{1\cdots}$), and sector-specific random effects. These consist of a varying intercept offset for work sector ($u_{0j}$) and a varying slope for sex within work sector ($u_{1j}\text{sex}_{i}$), where $j$ indexes which work sector individual $i$ belongs to. A simplified representation of the models is shown in Equation 2.

$$\begin{matrix} y_{i}\sim\text{Bernoulli}\left( p_{i} \right) \\ \text{logit}\left( p_{i} \right)=\alpha+\hat{\beta_{1}}*\text{sex}_{i}+\hat{\beta_{2}}*\text{age group}_{i}+\hat{\beta_{3}}*\text{ethnicity}_{i}+\cdots+u_{0j}+u_{1j}\text{sex}_{i} \end{matrix} \left( 2 \right)$$

Models M3 and M4 are two-part hurdle models, where $y$ is the number of days of presenteeism and can be zero or a positive count; $z$ is the probability of a structural zero and $f_{z}$ is the probability of zero in the underlying count distribution. Equation 3 describes the hurdle mechanism.

$$\begin{matrix} f_{z}\left( y_{i} \right) = z \text{if} y_{i}=0 \\ f_{z}\left( y_{i} \right) = \left( 1-z \right) f\left( y_{i} \right)/\left( 1-f\left( 0 \right) \right) \text{if} y_{i}>0 \\ \end{matrix} \left( 3 \right)$$

The hurdle component (see Equation 4) is estimated using a Bernouilli distribution and estimates whether the number of presenteeism days is zero. Here, $p_{i}$ is the probability that individual $i$ had a number of presenteeism days greater than 0 in the study period.

$$\begin{matrix} y_{i}\sim\text{Bernoulli}\left( p_{i} \right) \\ \text{logit}\left( p_{i} \right)=\alpha+\hat{\beta_{1}}*\text{sex}_{i}+\hat{\beta_{2}}*\text{age group}_{i}+\hat{\beta_{3}}*\text{ethnicity}_{i}+\cdots+u_{0j}+u_{1j}\text{sex}_{i} \end{matrix} \left( 4 \right)$$

The conditional (count) component, shown in Equation 5, estimates the number of presenteeism days using a negative binomial distribution where $\theta$ represents the dispersion parameter and $y$ is the number of days of presenteeism, which in this component must be a positive count

$$\begin{matrix} y_{i}\sim\text{NegativeBinomial}\left( \mu_{i},\theta\right) \\ \text{log}\left( \mu_{i} \right)=\alpha+\hat{\beta_{1}}*\text{sex}_{i}+\hat{\beta_{2}}*\text{age group}_{i}+\hat{\beta_{3}}*\text{ethnicity}_{i}+\cdots+u_{0j}+u_{1j}\text{sex}_{i} \end{matrix} \left( 5 \right)$$

M5 is an ordered Beta regression model. Ordered Beta models allow the modelling of ratio variables that also contain degenerate (0 or 1) values, by modelling a latent continuous variable and estimating two cutpoints that divide the distribution into zero values, values between zero and one, and one. Values between zero and one are modelled using a Beta distribution. This framework has been shown to be more accurate than alternatives such as zero-one-inflated Beta regression (Kubinec 2022).

The presenteeism rate $y_{i}$ for individual $i$ in work sector $j$ is calculated by dividing the number of presenteeism days of presenteeism for an individual by their number of potential work days. It is modelled as three sections: $y_{i}=0$ for no presenteeism, $0<y_{i}<1$ for some presenteeism, and $y_{i}=1$ for the individual working while sick with a respiratory infection during the entire period. We model $y_{i}$ using an ordered Beta regression with likelihood as shown in Equation 6:

$$f\left( y_{i}\mid X_{i},\beta_{i},\mu_{i},u_{0j},u_{1j}\text{sex}_{i},\phi,\kappa_{0},\kappa_{1} \right)=\left\{ \begin{matrix} 1-g\left( \eta_{i}-\kappa_{0} \right), & \text{if }y_{i}=0, \\ \left[ g\left( \eta_{i}-\kappa_{0} \right)-g\left( \eta_{i}-\kappa_{1} \right) \right]\cdot Beta\left( y_{i};\mu_{i},\phi\right), & \text{if }0<y_{i}<1, \\ g\left( \eta_{i}-\kappa_{1} \right), & \text{if }y_{i}=1. \end{matrix} \right. \left( 6 \right)$$

where $g\left( \cdot\right)$ is the inverse logit function, defined as:

$$g\left( x \right)=\frac{1}{1+e^{-x}} \left( 7 \right)$$

$\eta=X_{i}\beta$ is the linear predictor, and $\mu_{i}=g\left( \eta_{i} \right)$ is the mean of the Beta component. The Beta density $Beta\left( y_{i};\mu_{i},\phi\right)$ is parameterised in terms of the mean $\mu_{i}$ and the precision parameter $\phi$ with shape parameters $\alpha_{i}=\mu_{i}\phi$ and $\beta_{i}=\left( 1-\mu_{i} \right)\phi$. The cutpoints $k_{0}$ and $k_{1}$ separate the three regions and are constrained such that $k_{0}<k_{1}$, ensuring valid probabilities that sum to one.

##### Software and priors

Models M1 to M4 are estimated in the *brms* package (Bürkner 2017) and Models M5 and M6 were estimated in the *ordbetareg* package (Kubinec 2022), which is a custom interface to *brms.* In each case, the posterior was sampled with 4000 iterations and four chains, except for M5 which was sampled with 6000 iterations to increase the effective sample size. Convergence was checked by visual inspection of traceplots and by checking effective sample sizes.

Across all models, priors for intercepts($\alpha$) and fixed effects ($\beta$) were specified as Normal distributions with a mean of 0 and standard deviation of 1.5; estimating a priors-only model resulted in estimated 95% credible intervals for the odds ratio of fixed effects of 0.09 - 11.70, while the 50% credible intervals were around 0.36 - 2.75. Random effects priors were set as Student’s *t* distribution with 10 degrees of freedom, location of 0 and scale of 1.5. This resulted in prior distributions that were similar to those of the fixed effects, but with more regularization: the 95% credible intervals on the odds ratio scale were 0.07 - 14.42, and 50% credible intervals were 0.55 - 1.78. These priors are visualised in Supplementary Fig. 4. Note that the same priors are applied across all effects, i.e. bespoke priors are not specified for individual variables. Default *brms* LKJ(1) priors were used for random effect correlations.

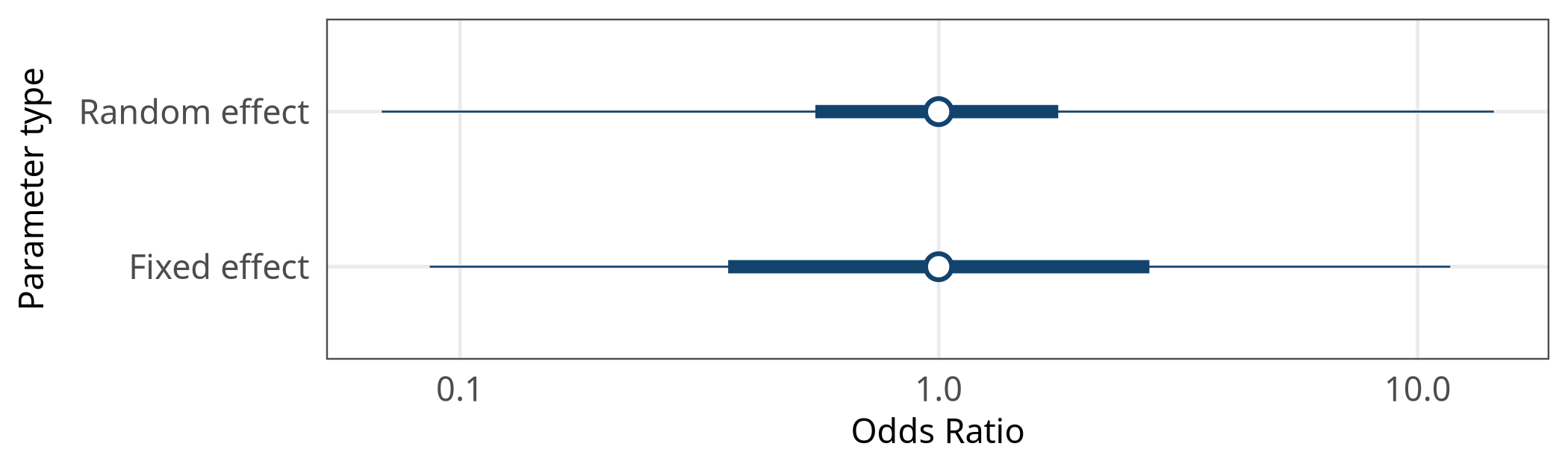

Supplementary Figure 4: Odds ratios with 50% and 95% credible intervals for model parameters, based only on the priors and not the data.

In the hurdle-negative binomial model, default *brms* priors were also used for dispersion (inverse Gamma with shape 0.4 and scale 0.3). In the ordered Beta regression models (M5 and M6), the default priors from the *ordbetareg* package were used for cutpoint locations ($k$) and the $\phi$ dispersion parameter. These are Dirichlet (1,1,1) priors and exponential (0.1) priors, respectively.

##### Model fits and posterior predictive checks

Model convergence statistics are shown in Supplementary Table 3. An Rhat of significantly above 1 indicates non-convergence, and an effective sample size of 1000 or above can be used as a rule of thumb to indicate stability (Bürkner 2017).

Supplementary Table 3: Convergence summary table showing the lowest Bulk Effective Sample Size (ESS), lowest Tail Effective Sample Size (ESS) and highest R-hat statistic from each model.

| Model | Max rhat | Min Bulk ESS | Min Tail ESS |
| --- | --- | --- | --- |
| M1 | 1.001703 | 1494.372 | 1763.442 |
| M2 | 1.002281 | 2459.628 | 2667.730 |
| M3 | 1.003760 | 1684.812 | 1674.042 |
| M4 | 1.001842 | 2022.647 | 1932.942 |
| M5 | 1.003234 | 2943.905 | 1855.218 |
| M6 | 1.002384 | 1941.984 | 2439.456 |

Figures Supplementary Fig. 5 to Supplementary Fig. 8 show the posterior predictive checks for models M1 to M6, comparing the actual data $y$ with simulated data from the posteriors $y_{rep}$. Models with a binary or count outcome variable are shown using bar plots, while density plots are used for models with continuous outcomes.

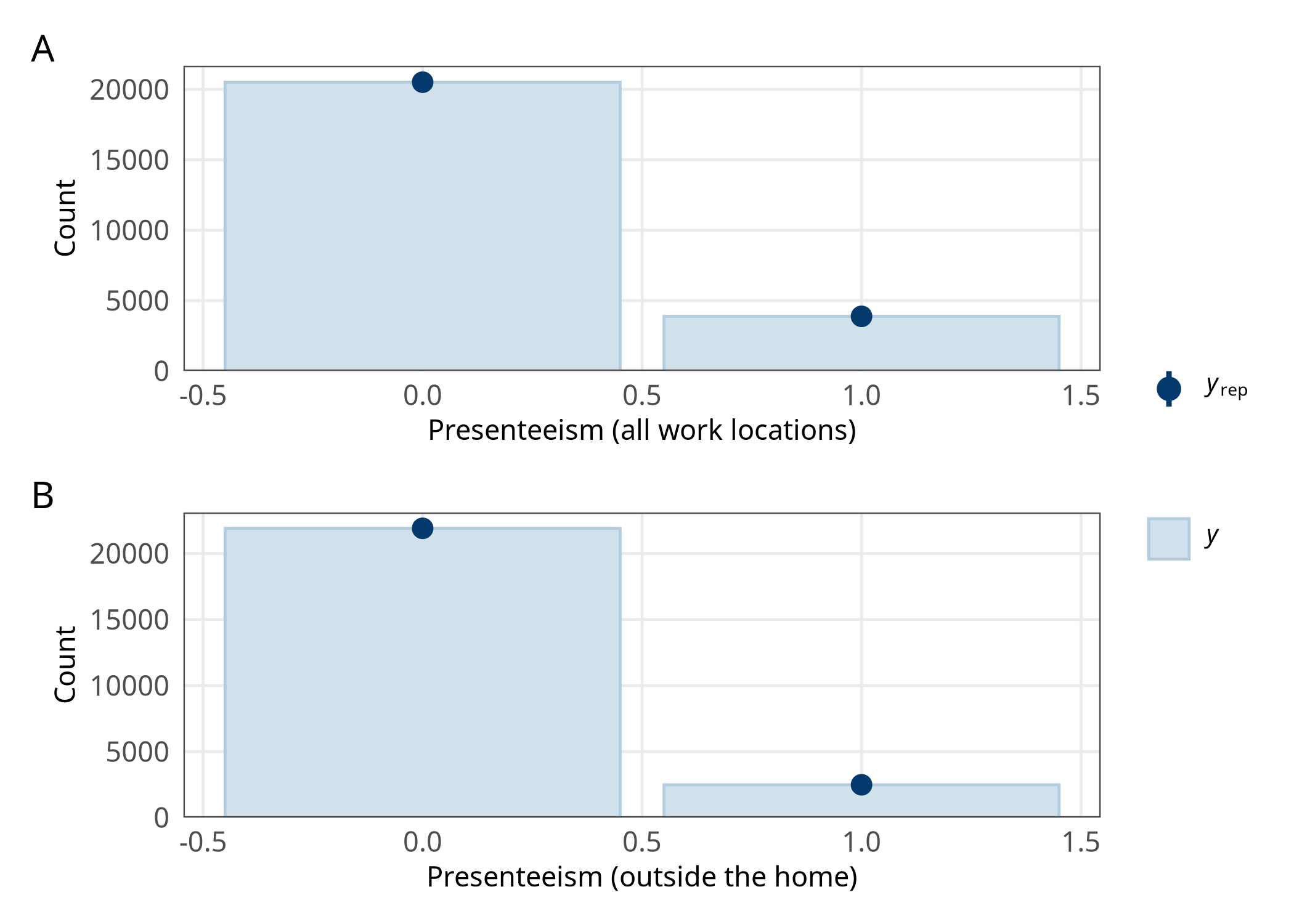

Supplementary Figure 5: Posterior predictive checks for models M1 (panel A) and M2 (panel B). These models estimate the proportion of working adults reporting presenteeism, in all work locations and outside the home.

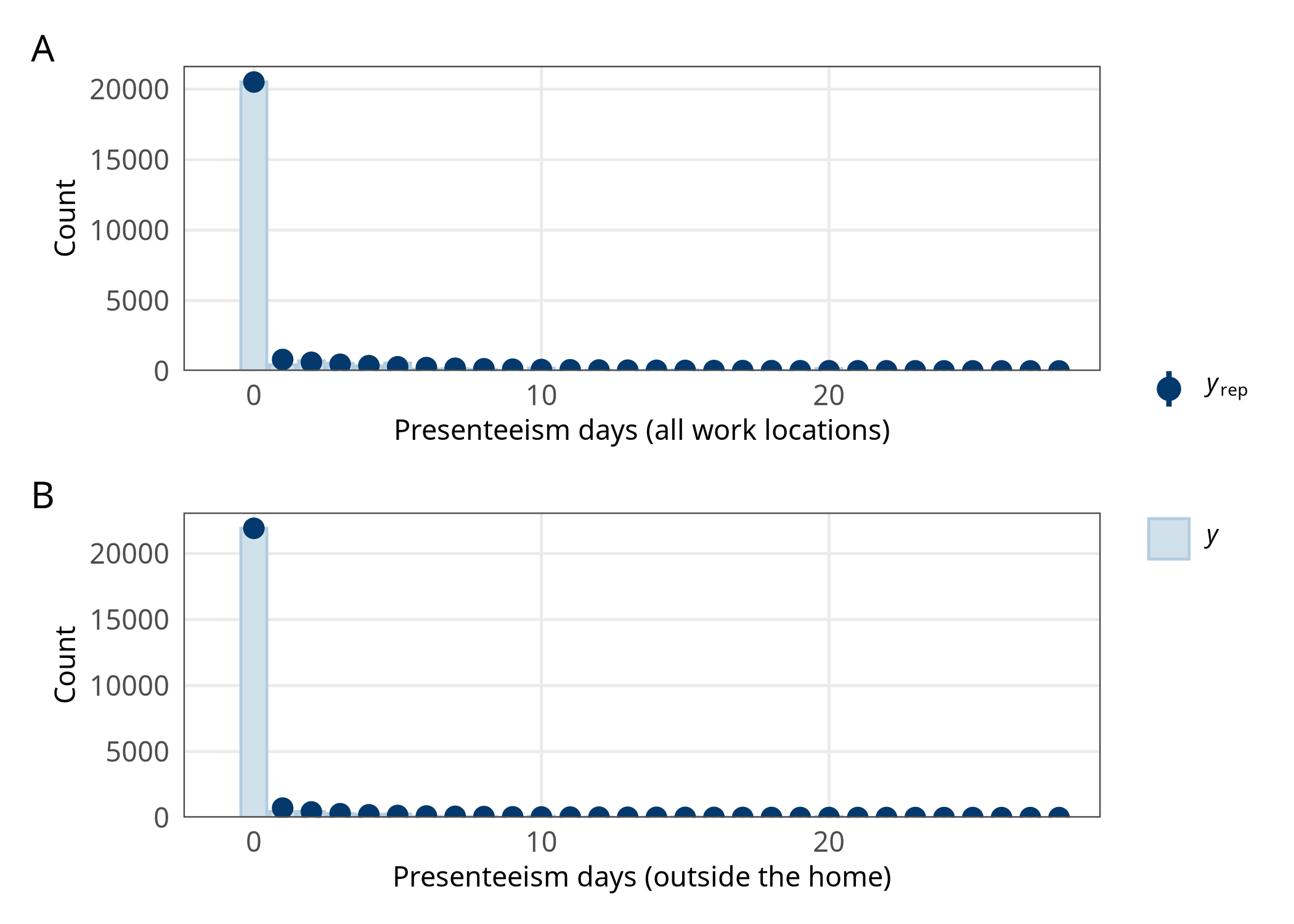

Supplementary Figure 6: Posterior predictive checks for models M3 and M4 (number of presenteeism days, overall and outside the home). X axis is truncated at 28 days to enhance visibility.

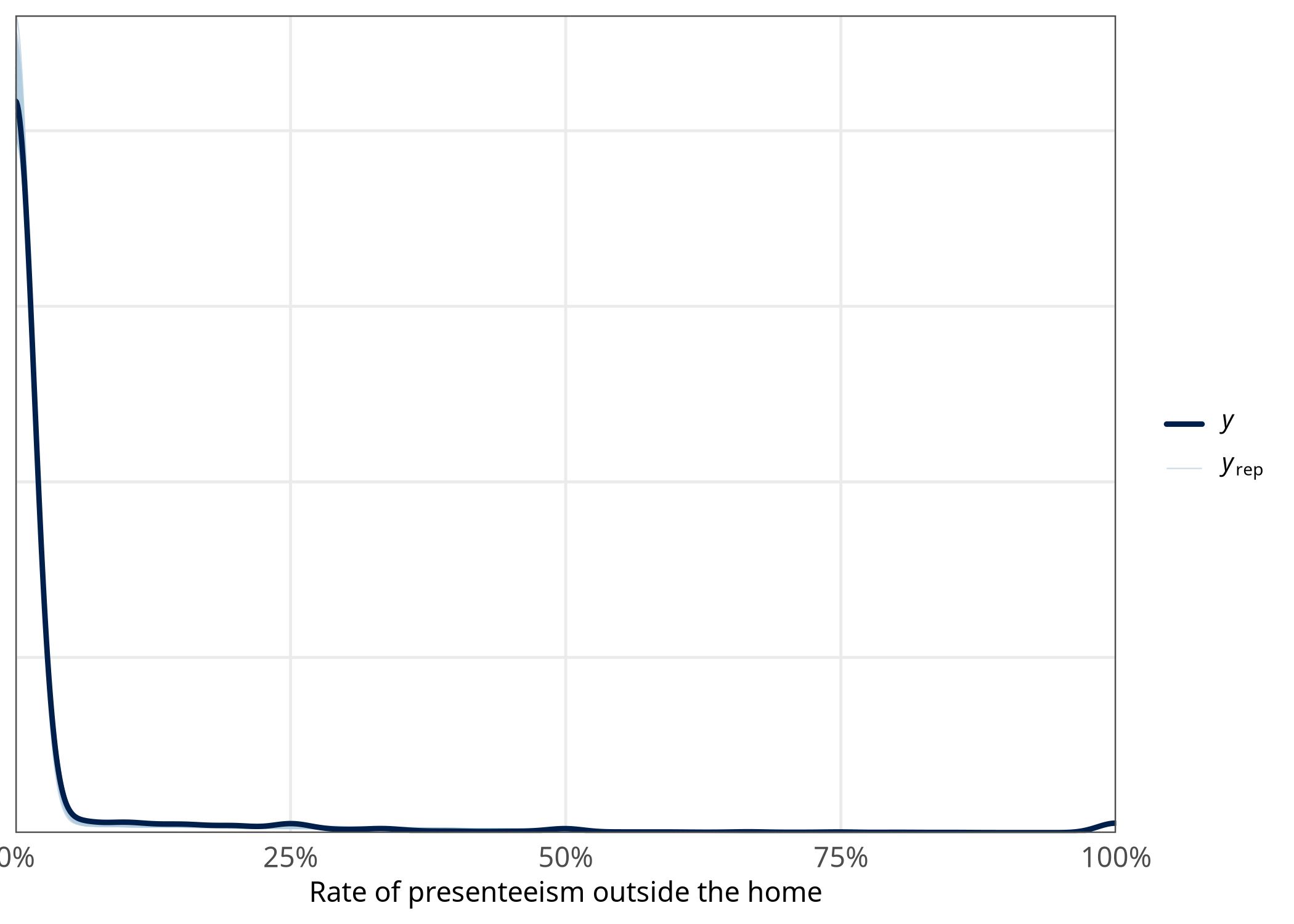

Supplementary Figure 7: Posterior predictive checks for model M5, which estimates the rate of presenteeism outside the home. Should be treated with caution, as it uses only the first imputed data set.

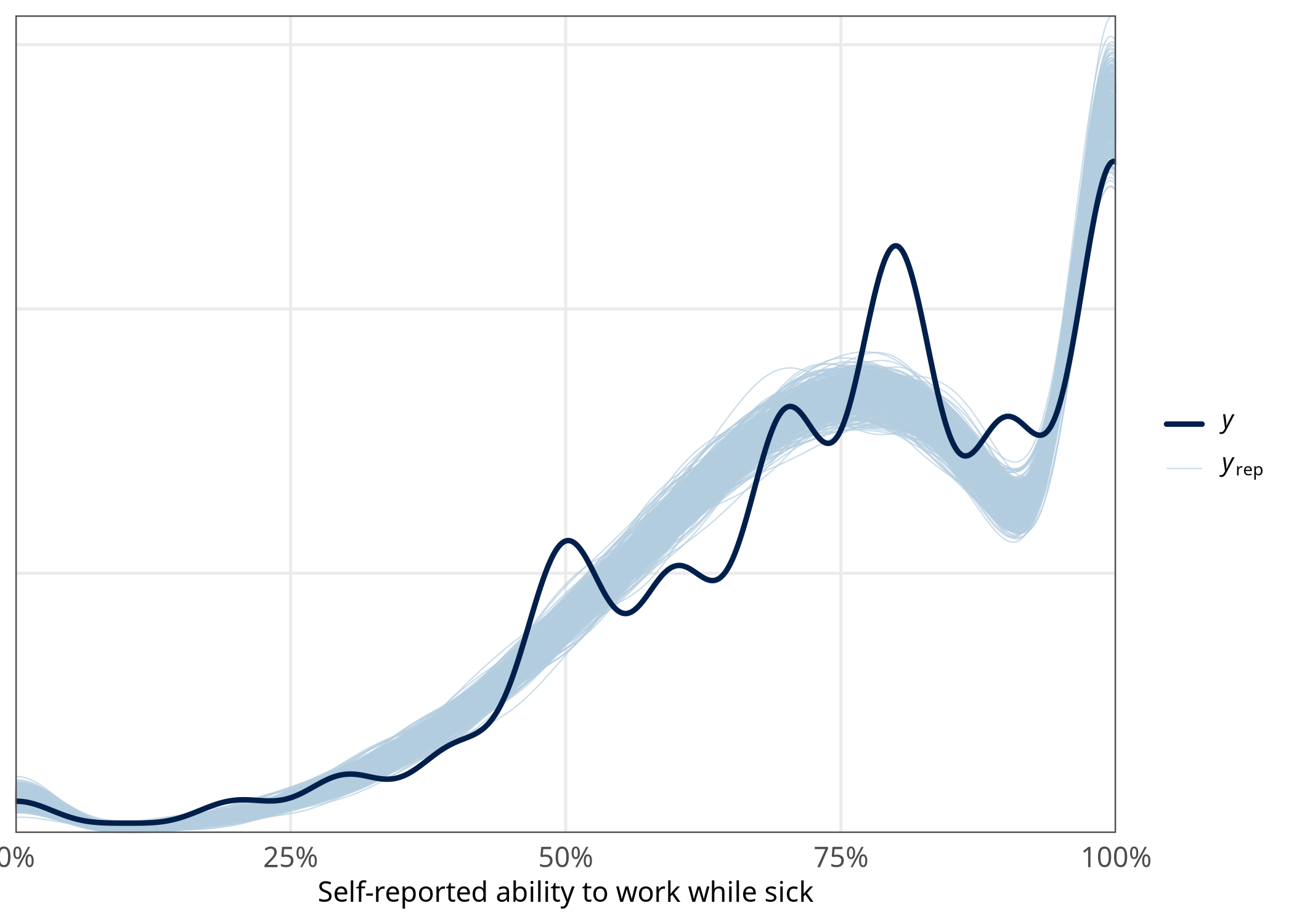

Supplementary Figure 8: Posterior predictive checks for model M6, which estimates self-reported ability to work while sick with a respiratory infection.

#### Results (average marginal effects tables)

##### Proportion of working adults who work while sick, overall and outside the home (M1 and M2)

Estimates of the proportion of working adults working while sick and the proportion of working adults working while sick outside the home with their 95% credible intervals are shown in Supplementary Table 4. These estimates are the average marginal effects of each variable and therefore represent changes in the estimated probability of presenteeism when moving between demographic groups, holding all other variables equal. To mitigate differences between the WCIS survey sample and known proportions, these estimates are poststratified by age group, sex, ethnicity and work sector. For categorical variables (e.g. work sector), the modal category was used as reference (age group 18-34, male, White, working outside the home, in the manufacturing or construction work sector).

Supplementary Table 4: Poststratified average marginal effects from the Bayesian logistic regression models M1 and M2, which model the proportion of working adults who work while sick (M1) or work outside the home while sick (M2). The reference category for each comparison is age group 18-34, male, White, working outside the home, in the manufacturing or construction work sector.

| Term | Poststratified AME [95% CrI] on presenteeism (from M1) | Poststratified AME [95% CrI] on presenteeism outside the home (from M2) |
| --- | --- | --- |
| Age group: 35-44 | 0.5% [-2.0%, 2.8%] | 1.4% [-0.5%, 3.3%] |
| Age group: 45-54 | -1.5% [-3.9%, 0.6%] | -0.1% [-1.9%, 1.6%] |
| Age group: 55-64 | -3.4% [-5.5%, -1.0%] | -1.4% [-3.3%, 0.3%] |
| Household size (+1) | 0.4% [-0.0%, 0.8%] | 0.2% [-0.1%, 0.5%] |
| IMD decile (+1) | 0.1% [-0.1%, 0.3%] | 0.1% [-0.1%, 0.3%] |
| Positive COVID LFT | 17.9% [11.6%, 25.8%] | 11.1% [5.5%, 17.2%] |
| Long COVID | 6.9% [4.2%, 9.5%] | 3.9% [2.0%, 6.1%] |
| Days of sickness absence (+1) | 7.8% [7.2%, 8.6%] | 3.1% [2.8%, 3.5%] |
| Region: East Midlands | 0.8% [-1.1%, 2.8%] | 1.0% [-0.6%, 2.7%] |
| Region: East of England | -0.3% [-2.0%, 1.5%] | 0.5% [-0.9%, 2.0%] |
| Region: North East | 1.3% [-1.2%, 3.9%] | 1.8% [-0.3%, 3.9%] |
| Region: North West | -0.6% [-2.4%, 1.1%] | 0.0% [-1.4%, 1.5%] |
| Region: South East | 0.3% [-1.4%, 1.8%] | 0.5% [-0.8%, 1.9%] |
| Region: South West | -0.1% [-1.9%, 1.6%] | -0.2% [-1.6%, 1.3%] |
| Region: West Midlands | 0.2% [-1.9%, 1.9%] | 1.0% [-0.6%, 2.6%] |
| Region: Yorkshire & the Humber | -0.5% [-2.4%, 1.4%] | 0.3% [-1.2%, 1.9%] |
| Female | 0.8% [-0.3%, 1.8%] | 0.4% [-0.4%, 1.3%] |
| Not White ethnicity | 3.3% [1.5%, 5.2%] | 2.7% [1.2%, 4.2%] |
| Work location: At home | -0.2% [-1.4%, 1.1%] | -12.1% [-13.2%, -11.1%] |
| Work location: Hybrid | 1.4% [0.2%, 2.5%] | -5.9% [-7.0%, -5.0%] |
| Work sector: Civil Service or Local Government & Armed forces | 1.8% [0.0%, 3.7%] | 1.1% [-0.5%, 2.8%] |
| Work sector: Financial services. This includes insurance | 0.0% [-1.8%, 1.8%] | -0.1% [-1.7%, 1.7%] |
| Work sector: Food production and agriculture combined with hospitality | 0.4% [-1.6%, 2.4%] | 0.2% [-1.5%, 2.1%] |
| Work sector: Healthcare & social care | 0.2% [-1.4%, 1.8%] | 0.6% [-0.9%, 2.1%] |
| Work sector: Information technology and communication | 1.8% [-0.3%, 4.0%] | -1.0% [-2.9%, 0.9%] |
| Work sector: Other employment sector | 0.8% [-0.6%, 2.5%] | 0.1% [-1.2%, 1.5%] |
| Work sector: Retail sector. This includes wholesale | 0.2% [-1.5%, 2.1%] | -0.2% [-1.8%, 1.4%] |
| Work sector: Teaching and education | 2.4% [0.9%, 4.3%] | 2.9% [1.4%, 4.5%] |
| Work sector: Transport. This includes storage and logistics | 0.5% [-1.8%, 2.8%] | -0.3% [-2.2%, 1.6%] |
| Model type | Bayesian logistic regression | Bayesian logistic regression |

##### Number of days of presenteeism, overall and outside the home (M3 and M4)

Supplementary Table 5 shows the poststratified average marginal effects and 95% credible intervals for each variable on the estimated number of days in the 28-day study period that working adults worked while sick with a respiratory infection, in total (from M3) and outside the home (from M4).

Supplementary Table 5: Poststratified average marginal effects from the Bayesian hurdle-negative binomial models M3 and M4, which estimate the number of days worked while sick with a respiratory infection (in total and outside the home).

| Term | Poststratified AME [95% CrI] on days of presenteeism (from M3) | Poststratified AME [95% CrI] on days of presenteeism outside the home (from M4) |
| --- | --- | --- |
| Age group: 35-44 | 0.14 [-0.05, 0.30] | 0.11 [-0.02, 0.25] |
| Age group: 45-54 | 0.16 [-0.01, 0.33] | 0.15 [0.03, 0.27] |
| Age group: 55-64 | 0.05 [-0.11, 0.21] | 0.06 [-0.06, 0.18] |
| Days of sickness absence (+1) | 0.44 [0.39, 0.48] | 0.15 [0.13, 0.17] |
| Female | -0.02 [-0.11, 0.07] | -0.02 [-0.09, 0.06] |
| Household size (+1) | 0.03 [-0.00, 0.06] | 0.00 [-0.02, 0.03] |
| IMD decile (+1) | 0.00 [-0.01, 0.02] | -0.00 [-0.01, 0.01] |
| Long COVID | 0.78 [0.51, 1.06] | 0.40 [0.21, 0.60] |
| Not White ethnicity | 0.06 [-0.11, 0.23] | 0.09 [-0.04, 0.22] |
| Positive COVID LFT | 0.76 [0.29, 1.31] | 0.65 [0.25, 1.17] |
| Region: East Midlands | 0.16 [0.00, 0.35] | 0.19 [0.06, 0.33] |
| Region: East of England | 0.07 [-0.07, 0.21] | 0.07 [-0.03, 0.18] |
| Region: North East | 0.36 [0.09, 0.63] | 0.25 [0.08, 0.43] |
| Region: North West | 0.02 [-0.12, 0.16] | 0.03 [-0.06, 0.14] |
| Region: South East | 0.10 [-0.02, 0.24] | 0.13 [0.03, 0.24] |
| Region: South West | 0.05 [-0.10, 0.19] | 0.04 [-0.06, 0.14] |
| Region: West Midlands | 0.11 [-0.05, 0.27] | 0.09 [-0.03, 0.21] |
| Region: Yorkshire & the Humber | 0.07 [-0.09, 0.22] | 0.11 [-0.01, 0.22] |
| Work location: At home | 0.02 [-0.09, 0.13] | -0.78 [-0.87, -0.70] |
| Work location: Hybrid | -0.01 [-0.11, 0.07] | -0.53 [-0.61, -0.46] |
| Work sector: Civil Service or Local Government & Armed forces | 0.06 [-0.11, 0.22] | -0.00 [-0.12, 0.14] |
| Work sector: Financial services. This includes insurance | -0.06 [-0.21, 0.10] | -0.07 [-0.22, 0.07] |
| Work sector: Food production and agriculture combined with hospitality | -0.00 [-0.20, 0.16] | -0.05 [-0.19, 0.11] |
| Work sector: Healthcare & social care | -0.14 [-0.30, 0.01] | -0.12 [-0.25, 0.01] |
| Work sector: Information technology and communication | 0.07 [-0.10, 0.26] | -0.11 [-0.26, 0.04] |
| Work sector: Other employment sector | -0.01 [-0.15, 0.13] | -0.04 [-0.16, 0.07] |
| Work sector: Retail sector. This includes wholesale | 0.02 [-0.16, 0.17] | -0.00 [-0.14, 0.14] |
| Work sector: Teaching and education | 0.08 [-0.08, 0.22] | 0.10 [-0.04, 0.22] |
| Work sector: Transport. This includes storage and logistics | 0.03 [-0.16, 0.24] | 0.01 [-0.15, 0.17] |
| Model type | Bayesian hurdle negative binomial regression | Bayesian hurdle negative binomial regression |

##### Rate of days of presenteeism (M5)

Supplementary Table 6: Poststratified average marginal effects from the Bayesian ordered Beta regression model M5, which estimates the rate of days worked while sick with a respiratory infection outside the home.

| Term | Poststratified AME [95% CrI] on rate of presenteeism outside the home |
| --- | --- |
| Age group: 35-44 | 0.4% [-0.3%, 1.0%] |
| Age group: 45-54 | 0.2% [-0.4%, 0.8%] |
| Age group: 55-64 | -0.0% [-0.6%, 0.6%] |
| IMD decile (+1) | 0.0% [-0.0%, 0.1%] |
| Positive COVID LFT | 1.8% [0.2%, 3.4%] |
| Female | 0.2% [-0.2%, 0.5%] |
| Not White ethnicity | 0.4% [-0.2%, 1.0%] |
| Work location: At home | -4.3% [-4.7%, -4.0%] |
| Work location: Hybrid | -2.3% [-2.7%, -2.0%] |
| Household size (+1) | 0.1% [-0.0%, 0.2%] |
| Long COVID | 1.5% [0.7%, 2.3%] |
| Days of sickness absence (+1) | 0.5% [0.5%, 0.6%] |
| Region: East Midlands | 0.5% [-0.1%, 1.1%] |
| Region: East of England | 0.4% [-0.1%, 1.0%] |
| Region: North East | 1.2% [0.4%, 2.0%] |
| Region: North West | 0.3% [-0.2%, 0.8%] |
| Region: South East | 0.5% [0.1%, 1.0%] |
| Region: South West | 0.2% [-0.3%, 0.7%] |
| Region: West Midlands | 0.6% [-0.0%, 1.1%] |
| Region: Yorkshire & the Humber | 0.5% [-0.1%, 1.0%] |
| Work sector: Civil Service or Local Government & Armed forces | 0.2% [-0.4%, 0.7%] |
| Work sector: Financial services. This includes insurance | -0.1% [-0.7%, 0.5%] |
| Work sector: Food production and agriculture combined with hospitality | -0.0% [-0.8%, 0.5%] |
| Work sector: Healthcare & social care | -0.0% [-0.5%, 0.5%] |
| Work sector: Information technology and communication | -0.2% [-1.0%, 0.4%] |
| Work sector: Other employment sector | 0.0% [-0.5%, 0.5%] |
| Work sector: Retail sector. This includes wholesale | 0.2% [-0.4%, 0.8%] |
| Work sector: Teaching and education | 0.5% [-0.0%, 1.0%] |
| Work sector: Transport. This includes storage and logistics | 0.0% [-0.6%, 0.8%] |
| Model type | Bayesian ordered Beta regression |

##### Effect of presenteeism on productivity (M6)

Supplementary Table 7: Poststratified average marginal effects from the Bayesian ordered Beta regression model M6, which estimates the self-reported impact of sickness on productivity. Only respondents who reported being sick with a respiratory infection were included.

| Term | Poststratified AME [95% CrI] on estimated productivity while sick |
| --- | --- |
| Age group: 35-44 | 0.6% [-2.1%, 3.0%] |
| Age group: 45-54 | 2.5% [0.1%, 4.9%] |
| Age group: 55-64 | 3.9% [1.6%, 6.4%] |
| Household size (+1) | -0.0% [-0.5%, 0.4%] |
| IMD decile (+1) | 0.2% [-0.0%, 0.4%] |
| Positive COVID LFT | -1.3% [-5.5%, 2.5%] |
| Long COVID | -5.9% [-8.1%, -3.8%] |
| Days of sickness absence (+1) | -3.1% [-3.4%, -2.8%] |
| Region: East Midlands | 2.7% [0.5%, 4.9%] |
| Region: East of England | -0.5% [-2.5%, 1.4%] |
| Region: North East | 0.5% [-2.6%, 3.4%] |
| Region: North West | -1.0% [-3.1%, 1.1%] |
| Region: South East | -0.1% [-1.8%, 1.8%] |
| Region: South West | -0.2% [-2.2%, 1.9%] |
| Region: West Midlands | -1.0% [-3.3%, 1.2%] |
| Region: Yorkshire & the Humber | 0.3% [-1.9%, 2.5%] |
| Female | -1.3% [-2.6%, -0.2%] |
| Not White ethnicity | 3.2% [0.6%, 5.7%] |
| Work location: At home | -1.6% [-3.1%, -0.0%] |
| Work location: Hybrid | -2.1% [-3.3%, -0.8%] |
| Work sector: Civil Service or Local Government & Armed forces | -0.5% [-2.4%, 1.3%] |
| Work sector: Financial services. This includes insurance | -0.3% [-2.2%, 1.7%] |
| Work sector: Food production and agriculture combined with hospitality | -0.4% [-2.9%, 1.7%] |
| Work sector: Healthcare & social care | -0.8% [-2.8%, 1.0%] |
| Work sector: Information technology and communication | 0.1% [-1.8%, 2.2%] |
| Work sector: Other employment sector | -0.5% [-2.3%, 1.1%] |
| Work sector: Retail sector. This includes wholesale | 0.1% [-2.0%, 2.0%] |
| Work sector: Teaching and education | -2.1% [-4.2%, -0.1%] |
| Work sector: Transport. This includes storage and logistics | -0.4% [-2.8%, 2.2%] |
| Model type | Bayesian ordered Beta regression |

Supplementary Table 8: Estimated productivity impacts of presenteeism by work sector, based on combining estimated days of presenteeism with self-reported ability to work while sick with a respiratory infection and Census estimates for numbers of adults working in each work sector. Modelled estimates and 95% highest density intervals are shown for the average days of productivity lost per person and years of productivity lost in total for each work sector.

| Work sector | Number of people | Productivity days lost per person | Total productivity years lost |
| --- | --- | --- | --- |
| Civil Service, Local Gov’t / Armed Forces | 1,436,247 | 0.24 [0.21, 0.28] | 963 [833, 1085] |
| Financial services | 983,319 | 0.21 [0.18, 0.24] | 561 [486, 650] |
| Food production & agriculture / Hospitality | 1,365,714 | 0.24 [0.20, 0.29] | 902 [736, 1082] |
| Health and social care | 3,680,442 | 0.18 [0.16, 0.21] | 1857 [1604, 2129] |
| IT and communication | 1,212,338 | 0.23 [0.20, 0.27] | 764 [653, 880] |
| Manufacturing or construction | 4,366,239 | 0.20 [0.17, 0.23] | 2411 [2082, 2749] |
| Other | 4,119,426 | 0.21 [0.19, 0.24] | 2397 [2129, 2657] |
| Retail | 4,095,985 | 0.21 [0.18, 0.25] | 2410 [2012, 2838] |
| Teaching and education | 2,483,662 | 0.29 [0.26, 0.32] | 1958 [1748, 2181] |
| Transport | 1,262,028 | 0.23 [0.19, 0.28] | 808 [663, 976] |

#### Results (raw model outputs)

### M1

| Term | Unweighted odds ratio [95% CrI] | Poststratified average marginal effect [95% CrI] |
| --- | --- | --- |
| (Intercept) | 0.15 [0.12 - 0.18] |  |
| Age group: 35-44 | 1.04 [0.87 - 1.24] | 0.48% [-1.95%, 2.86%] |
| Age group: 45-54 | 0.89 [0.75 - 1.05] | -1.50% [-3.92%, 0.65%] |
| Age group: 55-64 | 0.76 [0.64 - 0.9] | -3.41% [-5.67%, -1.17%] |
| Not White ethnicity | 0.76 [0.65 - 0.89] | 3.32% [1.38%, 5.06%] |
| Female | 1.07 [0.95 - 1.2] | 0.78% [-0.29%, 1.75%] |
| Positive COVID LFT | 2.8 [2.04 - 3.89] | 18.02% [11.56%, 25.53%] |
| Work location: At home | 0.99 [0.89 - 1.09] | -0.19% [-1.44%, 1.09%] |
| Work location: Hybrid | 1.11 [1.02 - 1.2] | 1.35% [0.23%, 2.47%] |
| IMD decile (+1) | 1.01 [1 - 1.02] | 0.13% [-0.05%, 0.32%] |
| Long COVID | 1.59 [1.36 - 1.86] | 6.88% [4.34%, 9.59%] |
| Household size (+1) | 1.03 [1 - 1.06] | 0.37% [-0.02%, 0.78%] |
| Days of sickness absence (+1) | 1.67 [1.61 - 1.74] | 7.82% [7.15%, 8.58%] |
| Region: East Midlands | 1.06 [0.92 - 1.24] | 0.82% [-1.06%, 2.90%] |
| Region: East of England | 0.98 [0.86 - 1.12] | -0.23% [-1.96%, 1.54%] |
| Region: North East | 1.1 [0.91 - 1.33] | 1.24% [-1.20%, 4.00%] |
| Region: North West | 0.95 [0.83 - 1.09] | -0.59% [-2.34%, 1.11%] |
| Region: South East | 1.02 [0.9 - 1.15] | 0.25% [-1.38%, 1.77%] |
| Region: South West | 0.99 [0.86 - 1.13] | -0.12% [-1.93%, 1.67%] |
| Region: West Midlands | 1.01 [0.87 - 1.17] | 0.12% [-1.70%, 2.16%] |
| Region: Yorkshire & the Humber | 0.96 [0.83 - 1.11] | -0.51% [-2.37%, 1.34%] |

The odds ratios and 95% credible intervals are also shown in Supplementary Fig. 9, as well as estimates for intercepts and slopes of the random effects.

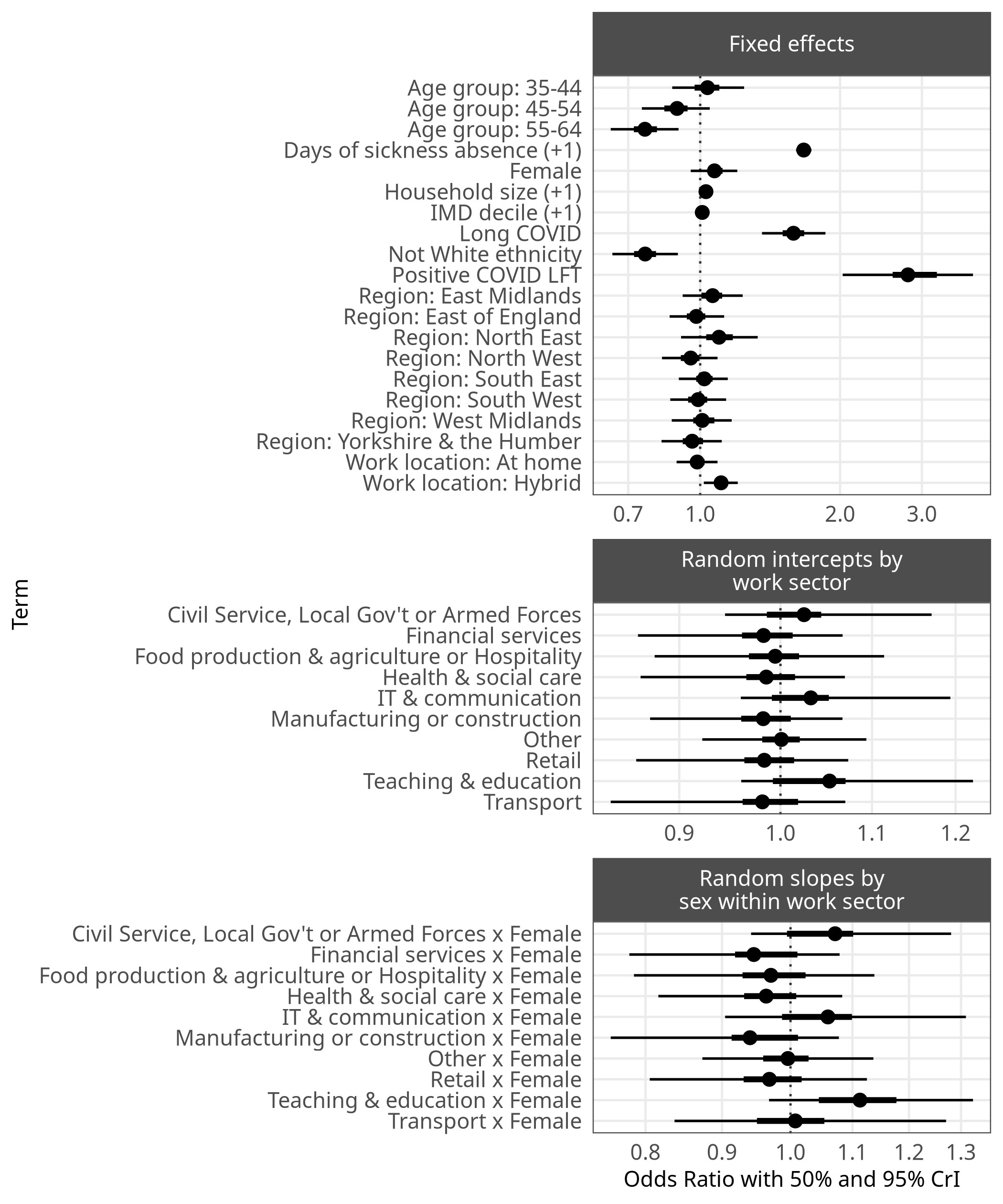

Supplementary Figure 9: Unweighted odds ratios with 50% and 95% credible intervals for effects on probability of working while sick from M1.

### M2

Supplementary Table 9: Unweighted odds ratios and poststratified average marginal effects from the Bayesian logistic regression model M2, which estimates effects on probability of working while sick outside the home.

| Term | Unweighted odds ratio [95% CrI] | Poststratified average marginal effect [95% CrI] |
| --- | --- | --- |
| (Intercept) | 0.13 [0.1 - 0.17] |  |
| Age group: 35-44 | 1.16 [0.93 - 1.43] | 1.33% [-0.58%, 3.30%] |
| Age group: 45-54 | 0.98 [0.81 - 1.21] | -0.15% [-1.91%, 1.64%] |
| Age group: 55-64 | 0.84 [0.69 - 1.04] | -1.42% [-3.12%, 0.36%] |
| Not White ethnicity | 0.71 [0.58 - 0.87] | 2.73% [1.20%, 4.19%] |
| Female | 1.04 [0.92 - 1.17] | 0.40% [-0.44%, 1.29%] |
| Positive COVID LFT | 2.51 [1.71 - 3.64] | 11.10% [5.50%, 17.17%] |
| Work location: At home | 0.12 [0.09 - 0.14] | -12.08% [-13.11%, -11.08%] |
| Work location: Hybrid | 0.53 [0.48 - 0.59] | -5.94% [-6.93%, -5.02%] |
| IMD decile (+1) | 1.01 [0.99 - 1.03] | 0.10% [-0.05%, 0.26%] |
| Long COVID | 1.48 [1.22 - 1.77] | 3.88% [1.76%, 5.90%] |
| Household size (+1) | 1.02 [0.98 - 1.06] | 0.16% [-0.15%, 0.49%] |
| Days of sickness absence (+1) | 1.37 [1.33 - 1.41] | 3.10% [2.77%, 3.50%] |
| Region: East Midlands | 1.12 [0.94 - 1.35] | 1.01% [-0.62%, 2.63%] |
| Region: East of England | 1.06 [0.89 - 1.24] | 0.51% [-0.92%, 1.91%] |
| Region: North East | 1.22 [0.97 - 1.52] | 1.80% [-0.24%, 3.88%] |
| Region: North West | 1 [0.84 - 1.18] | -0.02% [-1.38%, 1.47%] |
| Region: South East | 1.06 [0.92 - 1.24] | 0.54% [-0.71%, 1.85%] |
| Region: South West | 0.97 [0.81 - 1.15] | -0.27% [-1.61%, 1.24%] |
| Region: West Midlands | 1.12 [0.93 - 1.34] | 0.97% [-0.59%, 2.62%] |
| Region: Yorkshire & the Humber | 1.04 [0.86 - 1.23] | 0.28% [-1.26%, 1.82%] |

### M3

Supplementary Table 10 shows the unweighted rate ratios for both the hurdle and conditional components of the hurdle-negative binomial model (M3), as well as the poststratified average marginal effect for each coefficient, with their respective 95% credible intervals. This model estimates the raw number of days in the 28-day study period that working adults worked while sick. Note that the coefficients for the hurdle component represent the likelihood of the response varible being zero, i.e. the respondent not working while sick at all - so unlike in other models or the conditional component, higher hurdle rate ratio coefficients represent *lower risk of presenteeism*. In addition, note that this model does not take into account how many days were worked during the study period.

Supplementary Table 10: Unweighted rate ratios and poststratified average marginal effects from the Bayesian hurdle-negative binomial model M3, which estimates the number of days worked while sick.

| Term | Hurdle RR [95% CrI] | Conditional RR [95% CrI] | Poststratified average marginal effect [95% CrI] |
| --- | --- | --- | --- |
| (Intercept) | 7.83 [5.95 - 10.32] | 3.89 [2.79 - 5.35] |  |
| Age group: 35-44 | 0.86 [0.69 - 1.06] | 1.11 [0.87 - 1.41] | 13.69% [-5.00% - 30.32%] |
| Age group: 45-54 | 1.02 [0.83 - 1.24] | 1.42 [1.12 - 1.75] | 16.37% [-1.40% - 32.08%] |
| Age group: 55-64 | 1.19 [0.97 - 1.44] | 1.4 [1.12 - 1.76] | 5.41% [-11.74% - 20.79%] |
| Not White ethnicity | 1.41 [1.16 - 1.74] | 1.17 [0.92 - 1.47] | 6.16% [-10.89% - 22.53%] |
| Female | 0.96 [0.85 - 1.08] | 0.91 [0.79 - 1.07] | -1.77% [-10.77% - 7.44%] |
| Positive COVID LFT | 0.39 [0.27 - 0.57] | 1.04 [0.73 - 1.47] | 76.05% [31.27% - 132.92%] |
| Work location: At home | 8.71 [7.01 - 10.73] | 0.48 [0.38 - 0.62] | -78.01% [-86.49% - -69.95%] |
| Work location: Hybrid | 1.88 [1.69 - 2.07] | 0.57 [0.51 - 0.63] | -53.27% [-61.55% - -45.78%] |
| IMD decile (+1) | 0.99 [0.97 - 1.01] | 0.99 [0.97 - 1.01] | -0.01% [-1.16% - 1.14%] |
| Long COVID | 0.67 [0.56 - 0.8] | 1.31 [1.11 - 1.58] | 39.72% [20.95% - 59.90%] |
| Household size (+1) | 0.98 [0.95 - 1.02] | 0.99 [0.95 - 1.03] | 0.39% [-2.03% - 2.79%] |
| Days of sickness absence (+1) | 0.73 [0.71 - 0.75] | 0.97 [0.95 - 0.99] | 14.99% [12.78% - 17.36%] |
| Region: East Midlands | 0.89 [0.74 - 1.07] | 1.34 [1.1 - 1.64] | 15.63% [-0.37% - 33.89%] |
| Region: East of England | 0.94 [0.8 - 1.11] | 1.13 [0.95 - 1.36] | 7.09% [-6.50% - 21.47%] |
| Region: North East | 0.82 [0.66 - 1.02] | 1.36 [1.08 - 1.71] | 36.09% [10.60% - 63.78%] |
| Region: North West | 1 [0.84 - 1.19] | 1.09 [0.91 - 1.31] | 2.52% [-11.49% - 16.45%] |
| Region: South East | 0.94 [0.8 - 1.09] | 1.28 [1.08 - 1.5] | 10.13% [-2.13% - 23.77%] |
| Region: South West | 1.03 [0.87 - 1.22] | 1.14 [0.95 - 1.39] | 4.60% [-9.53% - 19.38%] |
| Region: West Midlands | 0.9 [0.74 - 1.07] | 1.1 [0.91 - 1.34] | 10.78% [-4.69% - 27.75%] |
| Region: Yorkshire & the Humber | 0.97 [0.81 - 1.16] | 1.25 [1.04 - 1.5] | 7.19% [-8.45% - 22.52%] |

Because the hurdle negative binomial model has two separate components, these are plotted separately. Supplementary Fig. 10 plots the odds ratios for the hurdle component and Supplementary Fig. 11 plots the rate ratios for the conditional component.

Odds ratios with 50% and 95% credible intervals for the hurdle component of M3 are plotted in Supplementary Fig. 10. These represent how strongly each model term is associated with the probability of not working while sick at all during the study period.

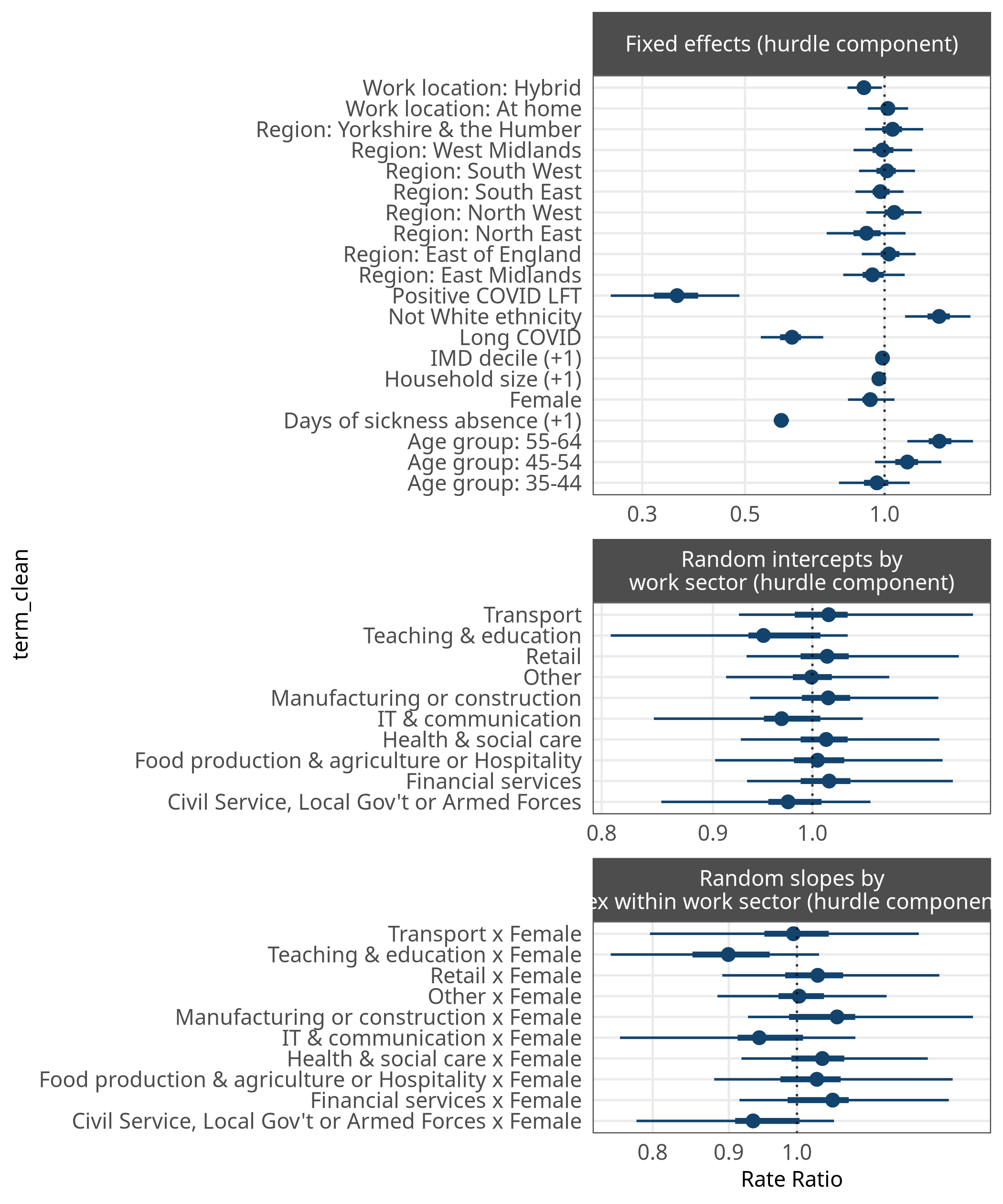

Supplementary Figure 10: Odds ratios with 50% and 95% credible intervals from the hurdle component of M3, indicating the association between each parameter and the likelihood of the response being zero (i.e., the respondent had no presenteeism in the study period).

Supplementary Fig. 11 plots the coefficients and 95% credible intervals for the conditional component of M3. These are rate ratios that represent the associations between each model term and the total number of days the respondent worked while sick, conditional on this being above zero.

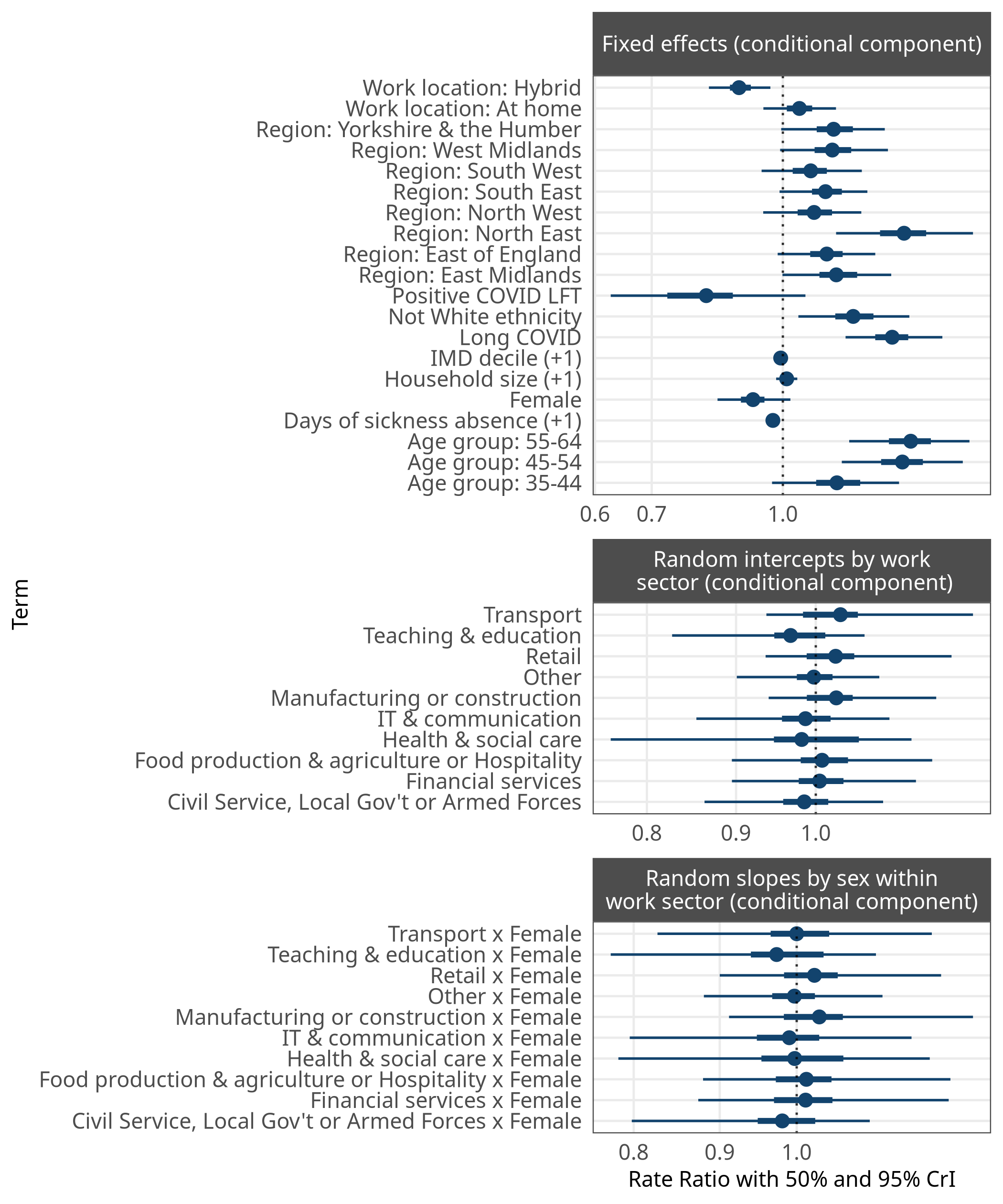

Supplementary Figure 11: Rate ratios with 50% and 95% credible intervals from the hurdle component of M3, indicating the association between each parameter and the likelihood of the response being zero (i.e., the respondent had no presenteeism in the study period).

### M4

Supplementary Table 11 shows the unweighted rate ratios for both the hurdle and conditional components of the hurdle-negative binomial model for presenteeism outside the home (M4), as well as the poststratified average marginal effect for each coefficient, with their respective 95% credible intervals. This model estimates the raw number of days in the 28-day study period that working adults worked while sick outside the home. Like M3, this model does not take into account how many days were worked during the study period.

Supplementary Table 11: Unweighted rate ratios and poststratified average marginal effects from the Bayesian hurdle-negative binomial model M4, which estimates the number of days worked while sick outside the home.

| Term | Hurdle RR [95% CrI] | Conditional RR [95% CrI] | Poststratified average marginal effect [95% CrI] |
| --- | --- | --- | --- |
| (Intercept) | 7.83 [5.95 - 10.32] | 3.89 [2.79 - 5.35] |  |
| Age group: 35-44 | 0.86 [0.69 - 1.06] | 1.11 [0.87 - 1.41] | 10.93% [-2.14%, 24.59%] |
| Age group: 45-54 | 1.02 [0.83 - 1.24] | 1.42 [1.12 - 1.75] | 14.73% [1.91%, 26.60%] |
| Age group: 55-64 | 1.19 [0.97 - 1.44] | 1.4 [1.12 - 1.76] | 6.15% [-7.05%, 17.27%] |
| Not White ethnicity | 1.41 [1.16 - 1.74] | 1.17 [0.92 - 1.47] | 9.13% [-5.06%, 20.56%] |
| Female | 0.96 [0.85 - 1.08] | 0.91 [0.79 - 1.07] | -1.76% [-9.03%, 5.15%] |
| Positive COVID LFT | 0.39 [0.27 - 0.57] | 1.04 [0.73 - 1.47] | 64.86% [25.85%, 118.30%] |
| Work location: At home | 8.71 [7.01 - 10.73] | 0.48 [0.38 - 0.62] | -78.01% [-86.49%, -69.95%] |
| Work location: Hybrid | 1.88 [1.69 - 2.07] | 0.57 [0.51 - 0.63] | -53.27% [-61.55%, -45.78%] |
| IMD decile (+1) | 0.99 [0.97 - 1.01] | 0.99 [0.97 - 1.01] | -0.01% [-1.16%, 1.14%] |
| Long COVID | 0.67 [0.56 - 0.8] | 1.31 [1.11 - 1.58] | 39.72% [20.95%, 59.90%] |
| Household size (+1) | 0.98 [0.95 - 1.02] | 0.99 [0.95 - 1.03] | 0.39% [-2.03%, 2.79%] |
| Days of sickness absence (+1) | 0.73 [0.71 - 0.75] | 0.97 [0.95 - 0.99] | 14.99% [12.78%, 17.36%] |
| Region: East Midlands | 0.89 [0.74 - 1.07] | 1.34 [1.1 - 1.64] | 18.60% [5.45%, 32.57%] |
| Region: East of England | 0.94 [0.8 - 1.11] | 1.13 [0.95 - 1.36] | 7.45% [-3.15%, 18.10%] |
| Region: North East | 0.82 [0.66 - 1.02] | 1.36 [1.08 - 1.71] | 24.46% [7.96%, 43.13%] |
| Region: North West | 1 [0.84 - 1.19] | 1.09 [0.91 - 1.31] | 3.36% [-6.45%, 13.85%] |
| Region: South East | 0.94 [0.8 - 1.09] | 1.28 [1.08 - 1.5] | 13.37% [3.35%, 23.78%] |
| Region: South West | 1.03 [0.87 - 1.22] | 1.14 [0.95 - 1.39] | 3.79% [-6.14%, 14.86%] |
| Region: West Midlands | 0.9 [0.74 - 1.07] | 1.1 [0.91 - 1.34] | 8.64% [-2.91%, 20.77%] |
| Region: Yorkshire & the Humber | 0.97 [0.81 - 1.16] | 1.25 [1.04 - 1.5] | 10.72% [-1.16%, 22.53%] |

Because the hurdle negative binomial model has two separate components, these are plotted separately. Supplementary Fig. 12 plots the odds ratios for the hurdle component and Supplementary Fig. 13 plots the rate ratios for the conditional component.

Odds ratios with 50% and 95% credible intervals for the hurdle component of M4 are plotted in Supplementary Fig. 12. These represent how strongly each model term is associated with the probability of not working outside the home while sick at all during the study period.

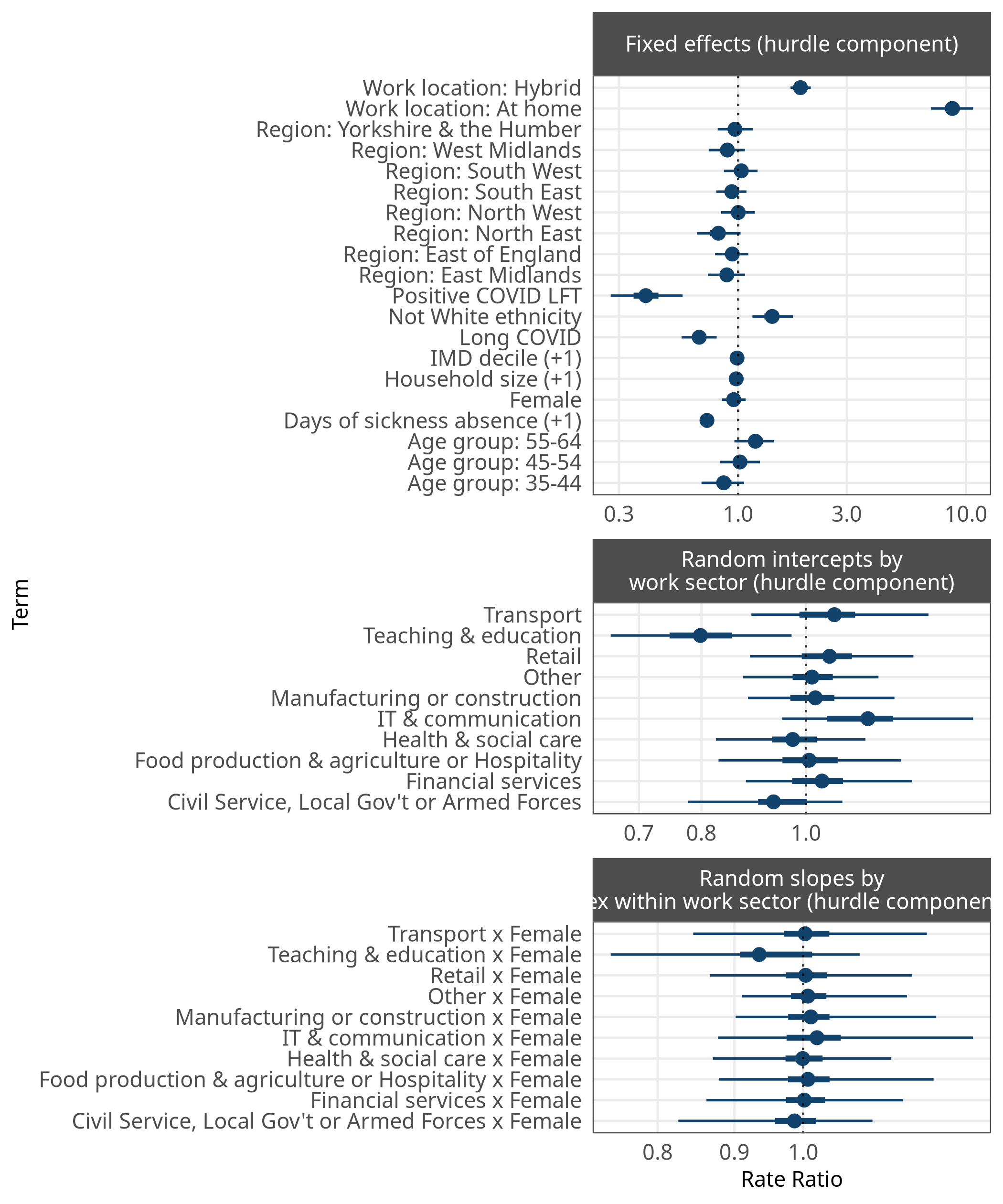

Supplementary Figure 12: Odds ratios with 50% and 95% credible intervals from the hurdle component of M4, indicating the association between each parameter and the likelihood of the response being zero (i.e., the respondent had no presenteeism outside the home in the study period).

Supplementary Fig. 13 plots the coefficients and 95% credible intervals for the conditional component of M4. These are rate ratios that represent the associations between each model term and the total number of days the respondent worked outside the home while sick, conditional on this being above zero.

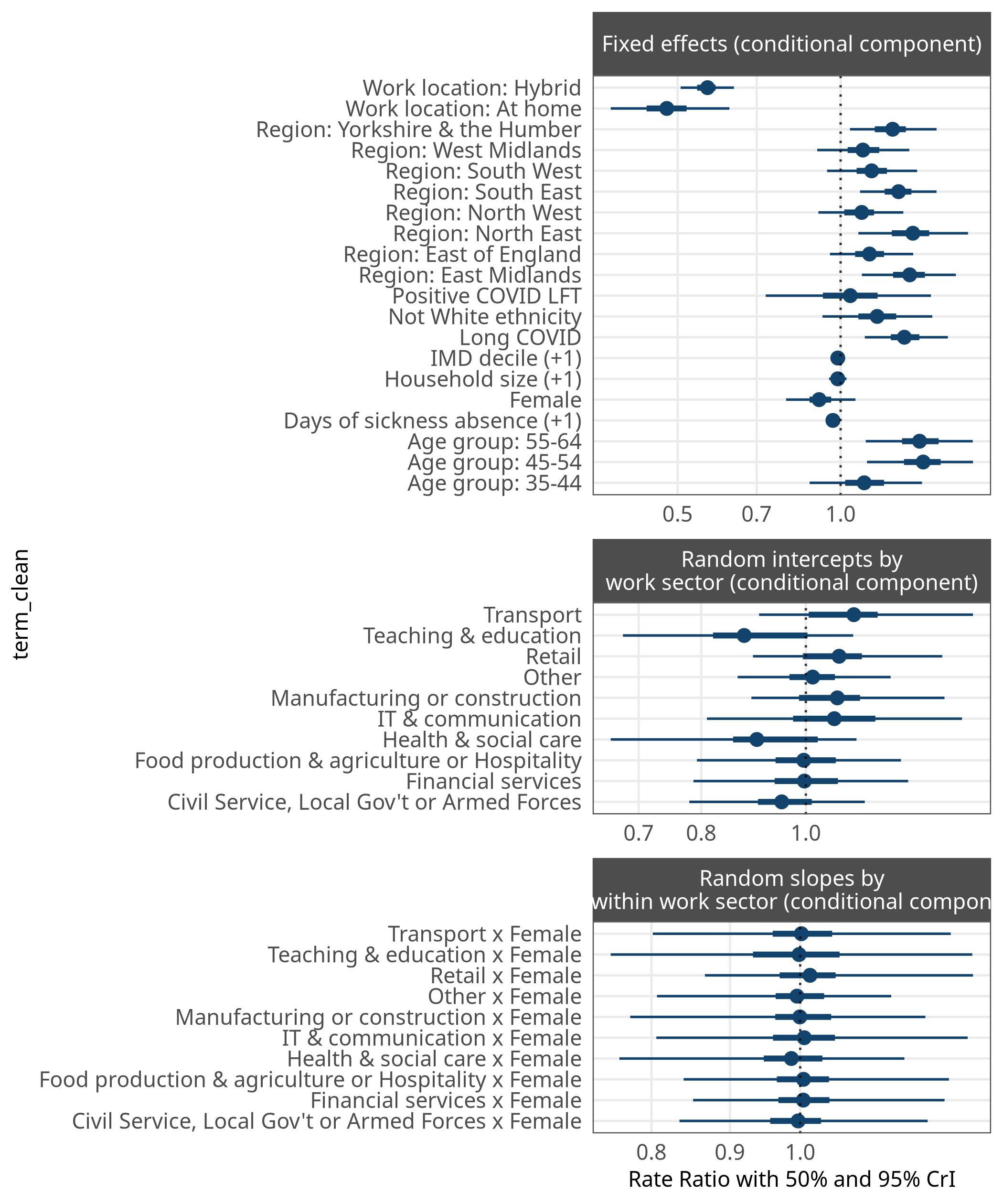

Supplementary Figure 13: Rate ratios with 50% and 95% credible intervals from the hurdle component of M4, indicating the association between each parameter and the likelihood of the response being zero (i.e., the respondent had no presenteeism outside the home in the study period).

### M5

| Term | Unweighted odds ratio [95% CrI] | Poststratified average marginal effect [95% CrI] |
| --- | --- | --- |
| (Intercept) | 0.3 [0.25 - 0.36] |  |
| Age group: 35-44 | 1.08 [0.94 - 1.24] | 0.39% [-0.37%, 1.01%] |
| Age group: 45-54 | 1.04 [0.91 - 1.18] | 0.17% [-0.45%, 0.79%] |
| Age group: 55-64 | 1 [0.88 - 1.14] | -0.01% [-0.62%, 0.60%] |
| Not White ethnicity | 0.91 [0.8 - 1.05] | 0.42% [-0.21%, 1.03%] |
| Female | 1.03 [0.94 - 1.12] | 0.15% [-0.17%, 0.48%] |
| Positive COVID LFT | 1.35 [1.07 - 1.69] | 1.77% [0.22%, 3.41%] |
| Work location: At home | 0.23 [0.2 - 0.27] | -4.34% [-4.72%, -3.96%] |
| Work location: Hybrid | 0.64 [0.6 - 0.69] | -2.30% [-2.64%, -1.94%] |
| IMD decile (+1) | 1.01 [1 - 1.02] | 0.03% [-0.02%, 0.09%] |
| Long COVID | 1.29 [1.15 - 1.45] | 1.47% [0.71%, 2.27%] |
| Household size (+1) | 1.02 [0.99 - 1.04] | 0.09% [-0.04%, 0.21%] |
| Days of sickness absence (+1) | 1.11 [1.09 - 1.13] | 0.54% [0.46%, 0.63%] |
| Region: East Midlands | 1.11 [0.98 - 1.26] | 0.50% [-0.09%, 1.09%] |
| Region: East of England | 1.1 [0.99 - 1.22] | 0.43% [-0.04%, 0.98%] |
| Region: North East | 1.26 [1.09 - 1.46] | 1.21% [0.43%, 2.08%] |
| Region: North West | 1.07 [0.96 - 1.2] | 0.34% [-0.17%, 0.84%] |
| Region: South East | 1.12 [1.02 - 1.24] | 0.54% [0.09%, 1.06%] |
| Region: South West | 1.05 [0.94 - 1.17] | 0.21% [-0.31%, 0.70%] |
| Region: West Midlands | 1.12 [1 - 1.26] | 0.54% [-0.02%, 1.12%] |
| Region: Yorkshire & the Humber | 1.1 [0.98 - 1.24] | 0.46% [-0.09%, 1.01%] |

The odds ratios and 95% credible intervals are also shown in Supplementary Fig. 14, as well as estimates for intercepts and slopes of the random effects.

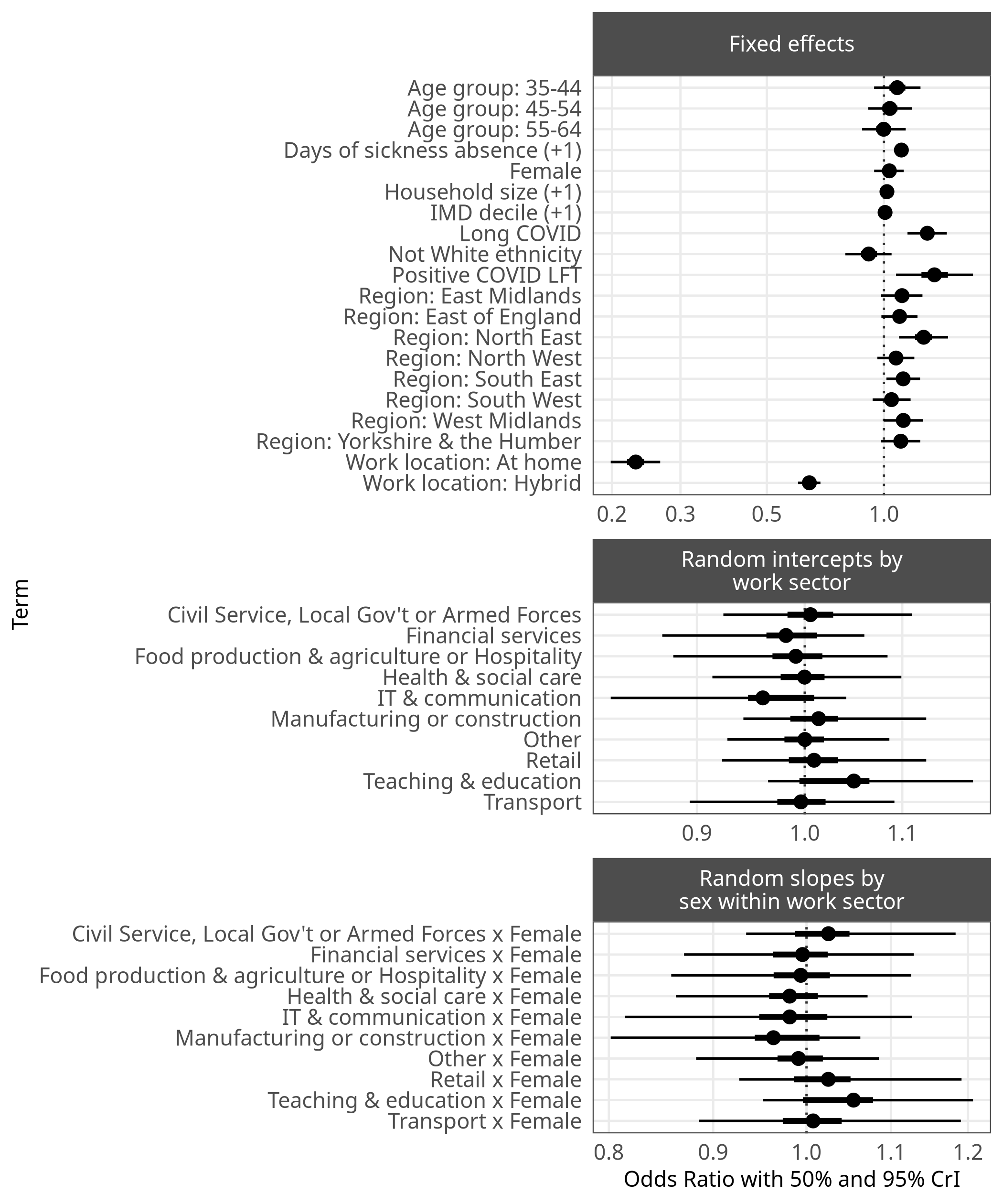

Supplementary Figure 14: Unweighted odds ratios with 50% and 95% credible intervals for the proportion of workdays worked while sick outside the home, using imputed data.

### M6

| Term | Unweighted odds ratio [95% CrI] | Poststratified average marginal effect [95% CrI] |
| --- | --- | --- |
| (Intercept) | 2.45 [2.12 - 2.85] |  |
| Age group: 35-44 | 1.03 [0.92 - 1.15] | 0.57% [-1.81%, 3.13%] |
| Age group: 45-54 | 1.12 [1.01 - 1.25] | 2.52% [0.15%, 4.76%] |
| Age group: 55-64 | 1.21 [1.08 - 1.34] | 3.99% [1.78%, 6.44%] |
| Not White ethnicity | 0.87 [0.78 - 0.97] | 3.14% [0.72%, 5.73%] |
| Female | 0.94 [0.87 - 1.02] | -1.38% [-2.61%, -0.11%] |
| Positive COVID LFT | 0.94 [0.8 - 1.13] | -1.35% [-5.07%, 2.54%] |
| Work location: At home | 0.93 [0.86 - 1] | -1.64% [-3.18%, -0.10%] |
| Work location: Hybrid | 0.91 [0.85 - 0.96] | -2.07% [-3.40%, -0.80%] |
| IMD decile (+1) | 1.01 [1 - 1.02] | 0.18% [-0.04%, 0.39%] |
| Long COVID | 0.77 [0.71 - 0.85] | -5.93% [-8.26%, -3.88%] |
| Household size (+1) | 1 [0.98 - 1.02] | -0.03% [-0.46%, 0.43%] |
| Days of sickness absence (+1) | 0.87 [0.86 - 0.88] | -3.07% [-3.37%, -2.80%] |
| Region: East Midlands | 1.14 [1.02 - 1.26] | 2.67% [0.58%, 4.89%] |
| Region: East of England | 0.98 [0.89 - 1.07] | -0.54% [-2.49%, 1.46%] |
| Region: North East | 1.02 [0.89 - 1.19] | 0.51% [-2.45%, 3.68%] |
| Region: North West | 0.96 [0.87 - 1.05] | -0.99% [-2.89%, 1.25%] |
| Region: South East | 1 [0.92 - 1.08] | -0.05% [-1.89%, 1.67%] |
| Region: South West | 0.99 [0.9 - 1.09] | -0.22% [-2.38%, 1.80%] |
| Region: West Midlands | 0.96 [0.86 - 1.06] | -0.95% [-3.19%, 1.41%] |
| Region: Yorkshire & the Humber | 1.01 [0.92 - 1.12] | 0.32% [-1.84%, 2.45%] |

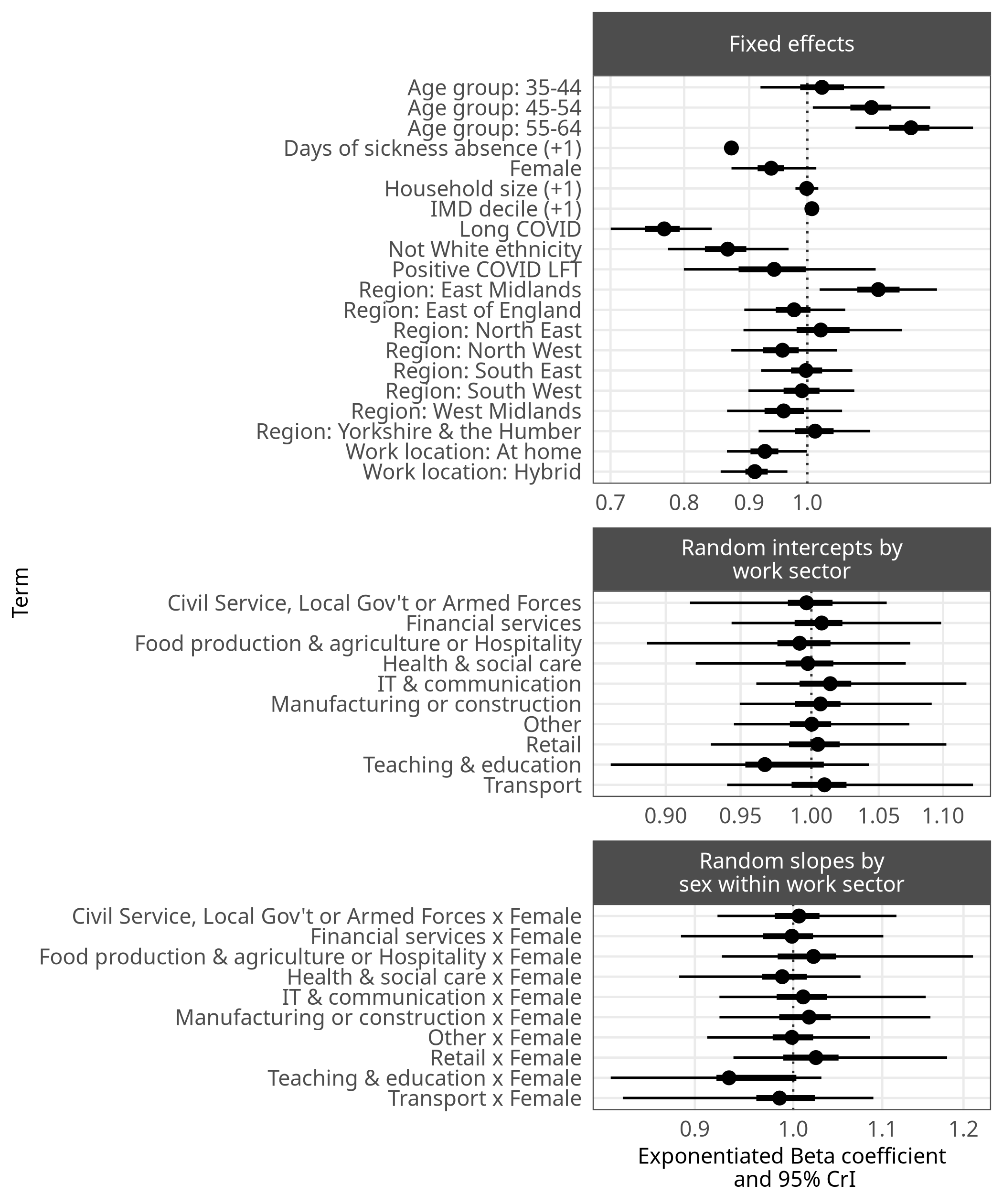

#### Sickness absence

To determine whether higher presenteeism in some groups was in addition to higher absenteeism, Supplementary Table 12 shows the poststratified average marginal effects of each variable on both days of presenteeism from model M3 and days sickness absence from model M3b,. Model M3b has the same specification as M3 but the response variable is number of days of sickness absence.

Supplementary Table 12: Poststratified average marginal effects of demographic variables on presenteeism and on sickness absence

| Term | Poststratified AME [95% CrI] on days of presenteeism (from M3) | Poststratified AME [95% CrI] on days of sickness absence (from M3b) |
| --- | --- | --- |
| Age group: 35-44 | 0.14 [-0.05, 0.30] | -0.04 [-0.10, 0.01] |
| Age group: 45-54 | 0.16 [-0.01, 0.33] | -0.02 [-0.08, 0.03] |
| Age group: 55-64 | 0.05 [-0.11, 0.21] | 0.01 [-0.05, 0.06] |
| Days of presenteeism (+1) | NA | 0.02 [0.02, 0.02] |
| Days of sickness absence (+1) | 0.44 [0.39, 0.48] | NA |
| Female | -0.02 [-0.11, 0.07] | 0.06 [0.04, 0.09] |
| Household size (+1) | 0.03 [-0.00, 0.06] | -0.00 [-0.01, 0.01] |
| IMD decile (+1) | 0.00 [-0.01, 0.02] | -0.01 [-0.01, -0.00] |
| Long COVID | 0.78 [0.51, 1.06] | 0.38 [0.26, 0.51] |
| Not White ethnicity | 0.06 [-0.11, 0.23] | 0.00 [-0.05, 0.05] |
| Positive COVID LFT | 0.76 [0.29, 1.31] | 0.41 [0.20, 0.67] |
| Region: East Midlands | 0.16 [0.00, 0.35] | -0.02 [-0.07, 0.03] |
| Region: East of England | 0.07 [-0.07, 0.21] | 0.01 [-0.03, 0.06] |
| Region: North East | 0.36 [0.09, 0.63] | -0.09 [-0.14, -0.05] |
| Region: North West | 0.02 [-0.12, 0.16] | -0.04 [-0.08, -0.00] |
| Region: South East | 0.10 [-0.02, 0.24] | 0.01 [-0.03, 0.05] |
| Region: South West | 0.05 [-0.10, 0.19] | 0.04 [-0.01, 0.10] |
| Region: West Midlands | 0.11 [-0.05, 0.27] | -0.02 [-0.06, 0.03] |
| Region: Yorkshire & the Humber | 0.07 [-0.09, 0.22] | -0.02 [-0.06, 0.03] |
| Work location: At home | 0.02 [-0.09, 0.13] | -0.03 [-0.06, 0.01] |
| Work location: Hybrid | -0.01 [-0.11, 0.07] | -0.02 [-0.05, 0.00] |
| Work sector: Civil Service or Local Government & Armed forces | 0.06 [-0.11, 0.22] | 0.03 [-0.01, 0.08] |
| Work sector: Financial services. This includes insurance | -0.06 [-0.21, 0.10] | 0.01 [-0.03, 0.06] |
| Work sector: Food production and agriculture combined with hospitality | -0.00 [-0.20, 0.16] | 0.01 [-0.04, 0.06] |
| Work sector: Healthcare & social care | -0.14 [-0.30, 0.01] | 0.03 [-0.01, 0.08] |
| Work sector: Information technology and communication | 0.07 [-0.10, 0.26] | 0.03 [-0.02, 0.08] |
| Work sector: Other employment sector | -0.01 [-0.15, 0.13] | 0.02 [-0.02, 0.06] |
| Work sector: Retail sector. This includes wholesale | 0.02 [-0.16, 0.17] | -0.02 [-0.06, 0.03] |
| Work sector: Teaching and education | 0.08 [-0.08, 0.22] | 0.04 [-0.01, 0.08] |
| Work sector: Transport. This includes storage and logistics | 0.03 [-0.16, 0.24] | 0.01 [-0.04, 0.07] |
| Model type | Bayesian hurdle negative binomial regression | Bayesian hurdle negative binomial regression |

#### Self-employment

As a supplement to the main analysis, in this section the role of self-employment is modelled. The WCIS data contained 2,972 self-employed respondents, which is 12% of the whole sample. In the raw data, 14% of self-employed respondents worked while sick during the study period, compared to 16% of employed respondents. We re-ran model M1 (likelihood of presenteeism) as M1a, including a binary variable for self-employment, to control for demographic differences between self-employed and employed respondents and to mitigate differences between the survey sample and the English working population.

Supplementary Table 13: Poststratified average marginal effects from the updated Bayesian logistic regression model M1a, which models the proportion of working adults who work while sick, but adds self-employment as a predictor. The reference category for each comparison is age group 18-34, male, White, working outside the home, in the manufacturing or construction work sector, and not self-employed.

| Term | Poststratified AME [95% CrI] on presenteeism (from M1a) |
| --- | --- |
| Age group: 35-44 | 0.5% [-1.7%, 2.9%] |
| Age group: 45-54 | -1.4% [-3.5%, 0.8%] |
| Age group: 55-64 | -3.1% [-5.3%, -1.0%] |
| Household size (+1) | 0.4% [-0.0%, 0.8%] |
| IMD decile (+1) | 0.1% [-0.1%, 0.3%] |
| Positive COVID LFT | 18.0% [11.2%, 25.0%] |
| Long COVID | 6.8% [4.2%, 9.4%] |
| Days of sickness absence (+1) | 7.8% [7.1%, 8.5%] |
| Region: East Midlands | 0.7% [-1.3%, 2.7%] |
| Region: East of England | -0.4% [-2.0%, 1.5%] |
| Region: North East | 1.1% [-1.5%, 3.7%] |
| Region: North West | -0.7% [-2.4%, 1.0%] |
| Region: South East | 0.2% [-1.4%, 1.7%] |
| Region: South West | -0.2% [-2.0%, 1.5%] |
| Region: West Midlands | 0.1% [-1.9%, 2.0%] |
| Region: Yorkshire & the Humber | -0.6% [-2.4%, 1.3%] |
| Female | 0.8% [-0.2%, 1.8%] |
| Not White ethnicity | 3.3% [1.4%, 5.0%] |
| Work location: At home | 0.2% [-1.2%, 1.5%] |
| Work location: Hybrid | 1.4% [0.4%, 2.6%] |
| Work sector: Civil Service or Local Government & Armed forces | 1.5% [-0.1%, 3.4%] |
| Work sector: Financial services. This includes insurance | -0.0% [-1.7%, 1.7%] |
| Work sector: Food production and agriculture combined with hospitality | 0.4% [-1.5%, 2.5%] |
| Work sector: Healthcare & social care | 0.2% [-1.4%, 1.8%] |
| Work sector: Information technology and communication | 1.7% [-0.3%, 3.9%] |
| Work sector: Other employment sector | 1.0% [-0.5%, 2.7%] |
| Work sector: Retail sector. This includes wholesale | 0.2% [-1.7%, 1.9%] |
| Work sector: Teaching and education | 2.3% [0.8%, 4.2%] |
| Work sector: Transport. This includes storage and logistics | 0.5% [-1.9%, 2.7%] |
| Self-employed | -2.7% [-4.1%, -1.3%] |
| Model type | Bayesian logistic regression |

As Supplementary Table 13 shows, on average self-employment was associated with a difference in probability of presenteeism of -2.7% (95% CI: -4.1%, -1.3%). This seems counter-intuitive, since employed individuals are more likely to be able to access paid sick leave than the self-employed. The result may be the result of having an unrepresentative sample of self-employed individuals, or the self-employed who work part-time may be better able to rearrange their workdays to accommodate sickness.
